# Plasma lipids and growth faltering: a longitudinal cohort study in rural Gambian children

**DOI:** 10.1101/2021.03.25.21253967

**Authors:** Gerard Bryan Gonzales, Daniella Brals, Bakary Sonko, Fatou Sosseh, Momodou Darboe, Andrew M. Prentice, Sophie E. Moore, Albert Koulman

## Abstract

Growth faltering in children arises from metabolic and endocrine dysfunction driven by complex interactions between poor diet, persistent infections and immunopathology. Here, we determined the progression of the plasma lipidome among Gambian children and assessed its influence on growth faltering over the first 2 years of life using panel vector autoregression modelling. We further investigated temporal associations among lipid clusters. We observed that measures of stunting, wasting and underweight significantly influence each other, and that lipid groups containing PUFA and phosphatidylcholines significantly influence growth outcomes. Linear growth was influenced by the majority of lipids, indicating a higher nutritional demand to improve height compared to weight among growth-restricted children. Our results indicate a critical role for PUFAs and choline in early life dietary interventions to combat the child growth faltering still so prevalent in low-income settings.

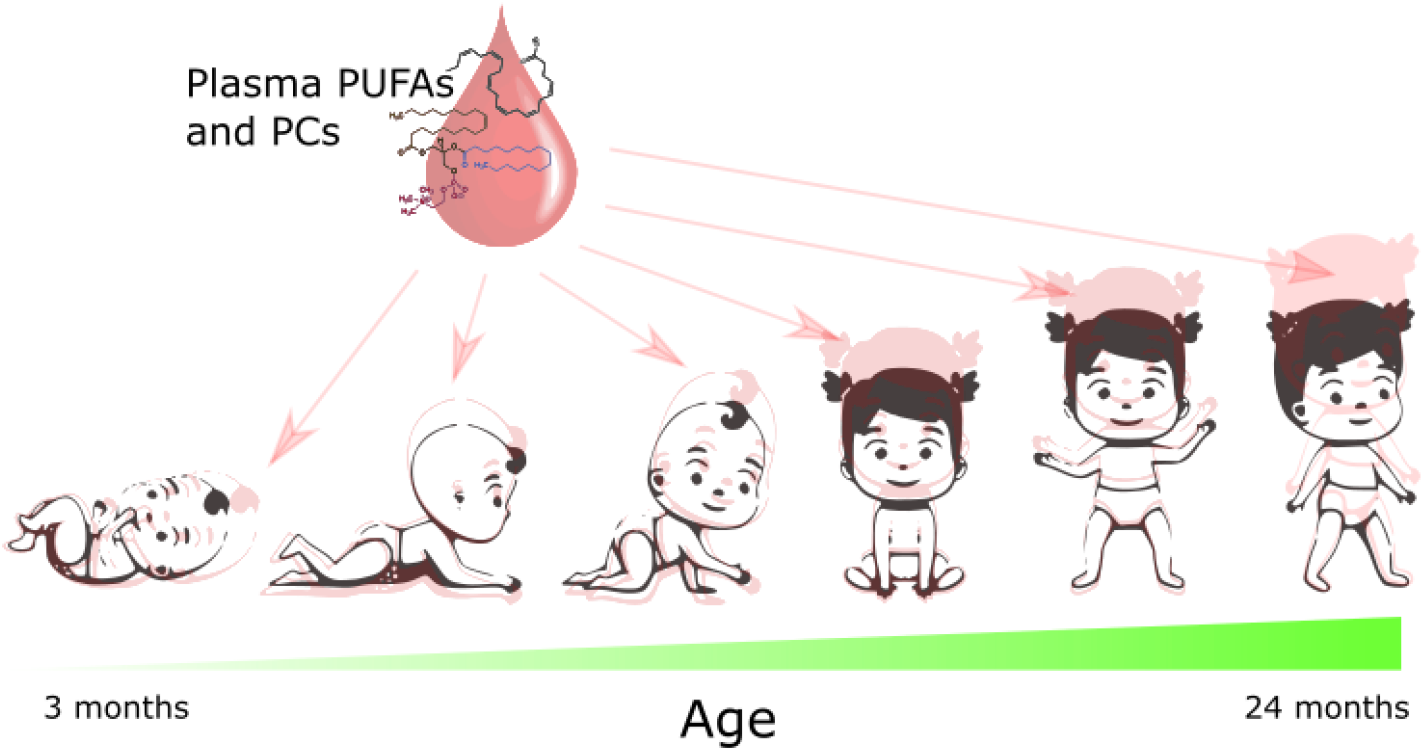

## Introduction

The first 1000 days (from conception to 2 years of age) are critically important in determining individual health trajectories to adulthood, and exposures during this period – especially nutritional exposures – can have lasting impact^1^. The causes of malnutrition are complex and multi-faceted, involving the interplay between nutrition, hygiene, infections, maternal health, economic status and other socio-demographic factors^2^. Malnutrition, which in this paper refers to undernutrition, is characterized by stunting (having a length-for-age z score (LAZ) below −2 standard deviations (SD)), wasting (below −2 SD weight-for-length (WLZ)) or underweight (below −2 SD weight-for-age (WAZ))^3^. Stunting is believed to be a result of chronic nutrient deprivation (chronic malnutrition), whereas wasting results from short-term malnutrition (hence, often referred to as acute malnutrition). Underweight is a reflection of both wasting and stunting^4^. Global estimates suggest that in 2019, 144 million children under 5 years of age were stunted while 47 million were wasted^5^, and this number is expected to rise due to the effect of the SARS-CoV-2 pandemic^6^.

Omics-based approaches have been used to gain a deeper understanding into the biochemical and metabolic perturbations that occur among children with malnutrition. However, the majority of these reports have focused on analysing samples and data from cross-sectional studies ^7-10^; data from longitudinal studies is needed to help understand the timing and direction of associations. By following the metabolome and lipidome progression over time in a single individual, resolution is enhanced, since inter-individual sources of variability (i.e. differences in (epi)genetic and lifestyle characteristics) are controlled. However, longitudinal analysis of high-dimensional data in field-based settings and among populations most at risk from undernutrition, especially metabolomics and lipidomics, remains challenging. Further, where longitudinal analyses exist, data analysis methods employed have been limited to assessing the progression of metabolic features over time, ranking the most dynamic features^11-15^, and not exploring potential causality or associations among the different metabolic features over time.

While the systems biology field has been exploring novel approaches to investigate longitudinal data and its association with specific clinical outcomes, other disciplines, such as econometrics and social sciences, have been analysing the same types of problems using robust data analysis approaches backed by strong mathematical foundations ^16-20^. In our current study, we investigated the association between longitudinal progression of the lipidome and growth outcomes in the first 2 years of life among children in The Gambia using an econometric approach applied to systems biology. Here, we adopt a panel vector autoregressive (PVAR) model in a generalized method of moments (GMM) framework to investigate the directions of potential causation between serum lipids and growth outcomes in these children.

## Results

### Population characteristics

A total of 1631 serum samples were analysed from 409 individual children from 3 months of age up to 2 years (5 time points). A total of 205 children had samples from all 5 timepoints, 77 from 4 timepoints, 63 from 3 time points and the remainder (65) had samples from 2 timepoints ^21^. Table 1 highlights child characteristics, by timepoint. In general, a decline over time in WLZ, LAZ and WAZ was observed, indicating growth faltering in this population. Males had significantly lower WLZ, LAZ and WLZ than females across the first 2 years, but their growth patterns were not different from each other (i.e. no interaction between sex and age was found, p = 0.70).

**Table 1.**
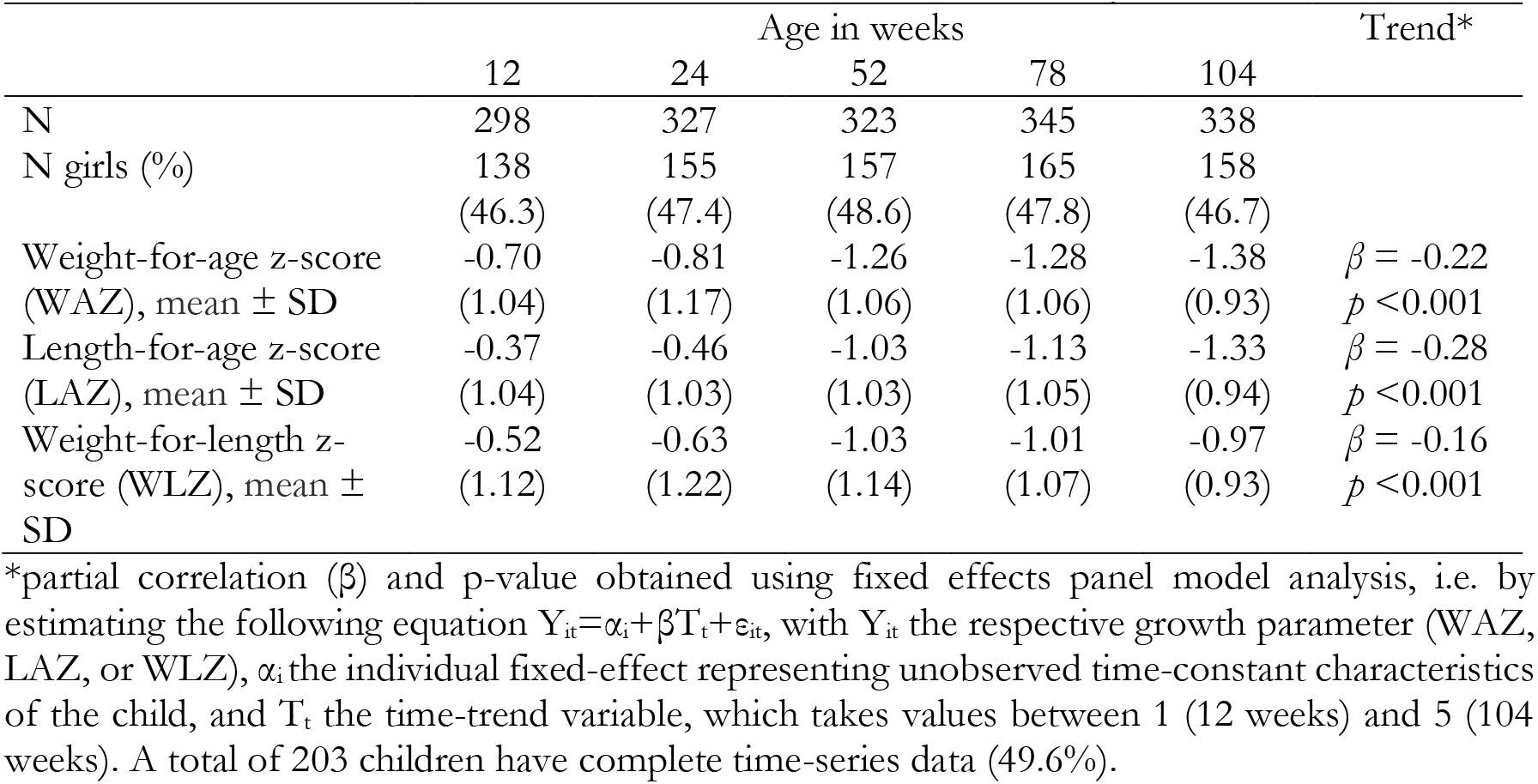
Growth characteristics of 410 Gambian children in the first 2 years of life

Using latent class linear mixed models, we identified sub-clusters within the population characterized by different growth patterns in the first 2 years of life. For LAZ, we identified 3 patterns of growth (Figure 1a). Cluster 1 (32%) included children who started with low LAZ at week 12 and maintained their LAZ over time. Cluster 2 (43%) included children with the highest LAZ at week 12, which gradually decreased over time but did not drop below −2 SD, indicating that children in this cluster were not considered stunted as classified by the World Health Organization (WHO) definition. Almost half (49.7%) of the children belonged to Cluster 3, which was characterized by mid-level LAZ at week 12 and having a steep decline in LAZ towards stunting over time. By week 104, 26% (25/95) of those in cluster 1 became stunted, whereas this was 47% (65/138) in cluster 3.

**Figure 1.**
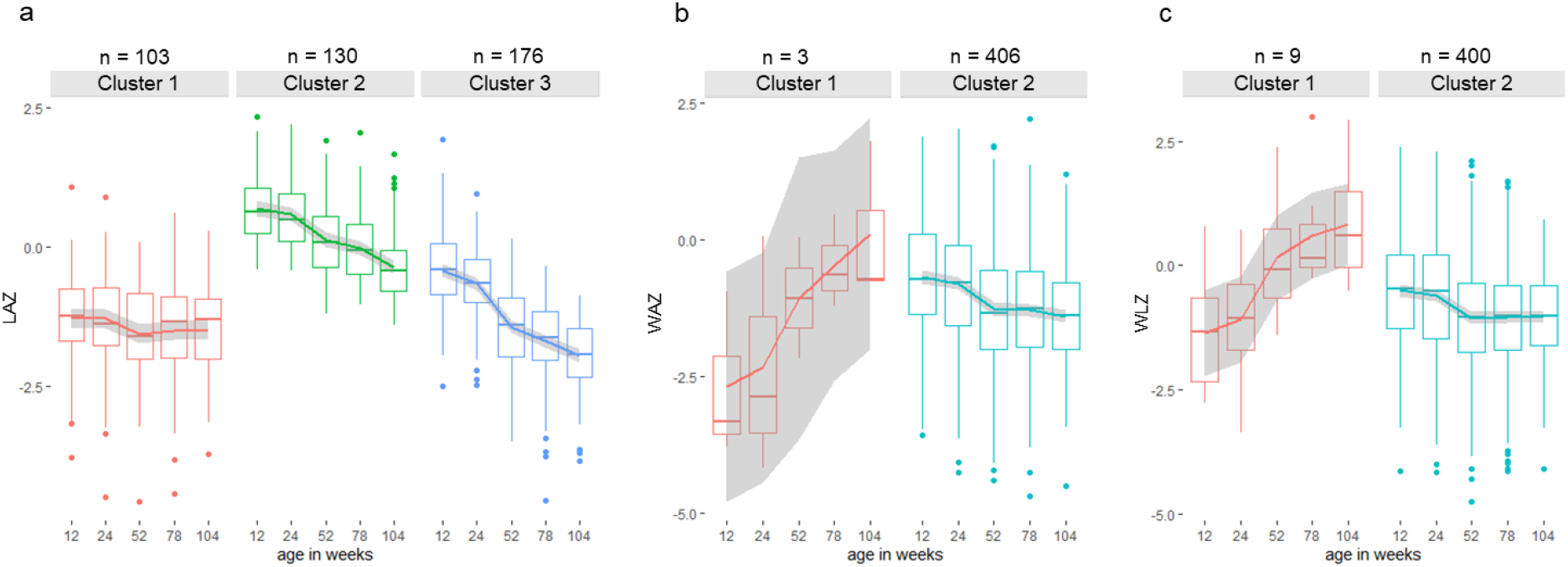
Growth patterns of children from 12 weeks to 104 weeks of life. Clusters of similar growth curves were generated using latent class mixed modelling. (a) 3 latent groups representing different LAZ progression in the population – 25% belonged to cluster 1, 32% to cluster 2, and 43% to cluster 3. For WAZ (b) and WLZ (c), 2 latent groups were obtained but 98-99% of the population belonged to cluster 2. A few children showed an increasing trend in their WAZ (3) and WLZ (9) in the first 2 years of life.

For WAZ (Figure 1b) and WLZ (Figure 1c), 2 clusters were identified but 98% and 99% of the children belonged to the second cluster for WAZ and WLZ, respectively. Three children had WAZ of −2.68 ± 1.52) at week 12 but caught-up in weight by week 104 (WAZ = 0.12 ± 1.47). In addition, the WLZ of 9 children at week 12 (WLZ = −1.36 ± 1.20) had significantly increased by week 104 (WHZ = 0.82 ± 1.13) (p < 0.05).

### Lipidome progression in the first 2 years of life

The total serum lipids (sum of all individual lipids) did not significantly change in the first 2 years of life, indicating that the lipid pool is conserved during infant growth (Figure 2a). However, serum lipid composition appeared to change over time. The serum concentration of most lipids identified (175/278, 63%) significantly decreased over time, whereas 17% (48/278) had a significant upward trend. Several lipids (55/278, 30%) on the other hand, were conserved during the first 2 years of life (Figure 2b). The progression of all identified lipids with age is shown in Supplementary Table 1.

**Figure 2.**
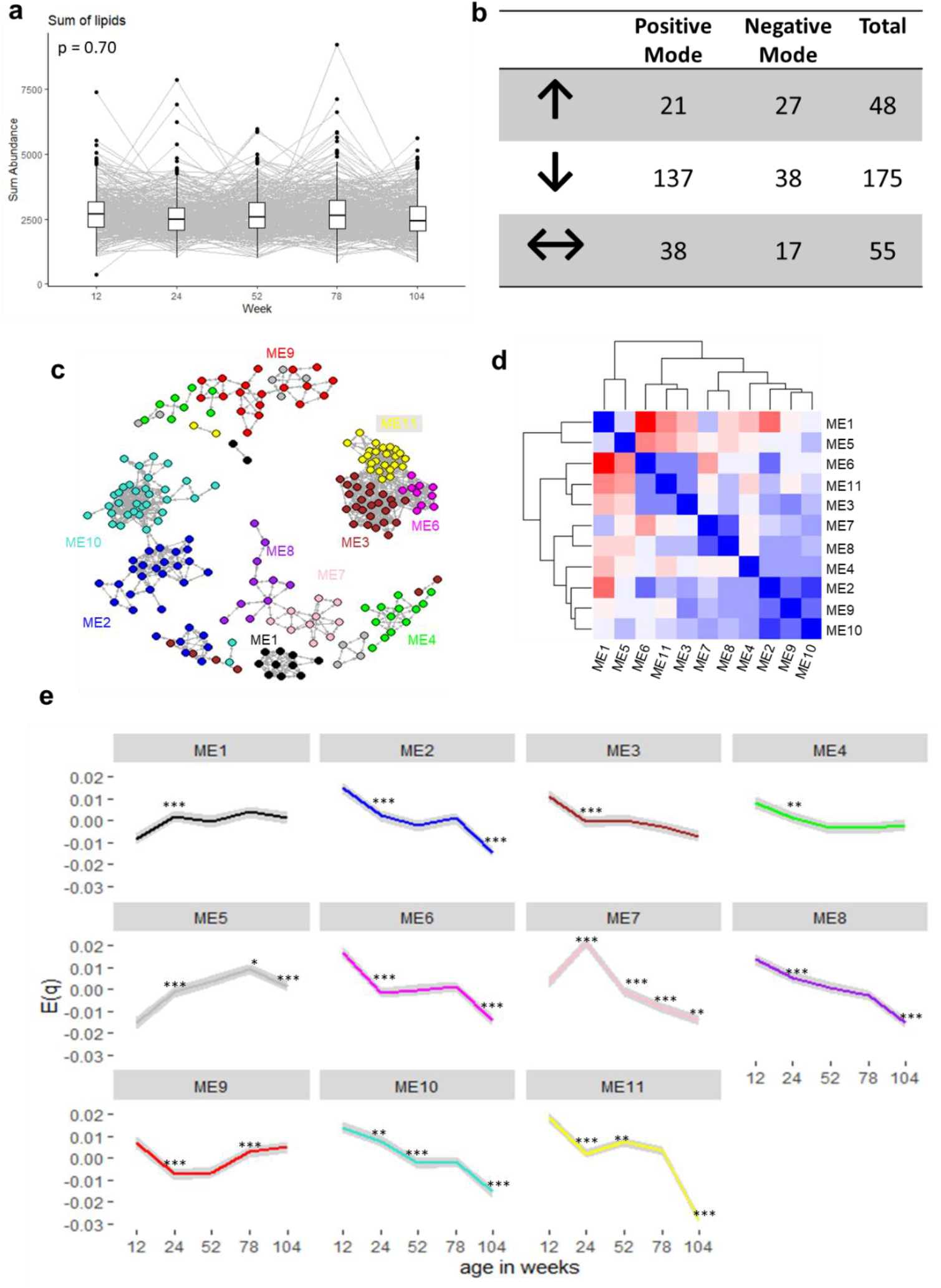
Lipid progression in the first 2 years of life among children in The Gambia. (**a**) Sum of total lipids over time. Fixed effects panel model revealed no significant change in total lipids through time (p = 0.70). (**b**) Number of lipids significantly (p = 0.05 adjusted for false discovery rate) altering through time; ↑ indicates significant increase, ↓ significant decrease and ↔ no significant change after Bonferonni correction. (**c**) Weighted correlation network showing 11 lipid clusters obtained using the WGCNA package in R. (**d**) Inter-modular relationship showing closely related lipid clusters. (**e**) Progression of eigenlipid (MEq, where q is the module number), which represents the collective behaviour of the lipids in the module, through time. Significance levels: *** indicate p < 0.0001, ** = p < 0.001, * = p <0.01. Comparisons were made using paired t-test comparing the time point and the preceding time point. Grey shadow for each line graph indicates the 95 % confidence interval.

To reduce the number of independent variables for succeeding analyses, we identified clusters of highly correlated lipids (“modules”) using weighted correlation network analysis^22^. Lipids assigned to their respective modules potentially share similar physiological and molecular characteristics, as modules reflect functional relationships (physical and non-physical interactions) among its members^22^. Each module is characterized by an eigenlipid (MEq, where q denotes the module), which is a unique representation that most closely reflects the collective behaviour of the module^23^. This indicates that the progression of lipids in each module through time is reflected by the dynamics of the MEq. About 87% (241/287) of the lipids were clustered into ten modules, whereas the remaining 37 (13%) lipid species were unassigned (grey module, ME5). Module assignment of all lipids are detailed in Supplementary Table 2. To obtain an overview of the inter-lipid correlations, we plotted the module correlation network (Figure 2c), which shows that several modules are more closely correlated, creating bigger clusters of lipids as depicted on a heatmap showing hierarchical clustering (Figure 2d).

**Table 2.** Association* between module eigenlipid (MEq) and growth outcomes through time in the first 2 years of life

| Module | Size<sup>§</sup> | Outcomes |  |  | Main Composition<sup>@</sup> |
| --- | --- | --- | --- | --- | --- |
|  |  | WAZ | LAZ | WLZ |  |
| ME1 | 16 | -2.65<br>($<0.001$) | -0.06<br>(0.92) | -3.67<br>($<0.0001$) | Ether-linked PCs and PSc, oxidised PCs |
| ME2 | 39 | 2.55<br>($<0.001$) | 0.58<br>(0.54) | 2.66<br>(0.011) | All PUFA-containing lipids (both n-3 (22:6, 20:5) and n-6 (20:4)), the cholesterol esters, PCs, PC-O/PC-PE-O/PE-P |
| ME3 | 31 | -1.36<br>(0.14) | -1.47<br>(0.19) | -1.49<br>(0.22) | Most common TGs and DGs |
| ME4 | 26 | 0.13<br>(0.84) | -0.08<br>(0.92) | 0.47<br>(0.70) | PA, PEs |
| ME5 | 37 | -2.43<br>($<0.001$) | -0.34<br>(0.71) | -3.67<br>($<0.0001$) | Unassigned lipids; free FAs and FA oxidation products and their esters |
| ME6 | 11 | 1.26<br>(0.15) | -0.59<br>(0.54) | 1.41<br>(0.22) | TG containing PUFAs |
| ME7 | 14 | 0.26<br>(0.74) | 0.76<br>(0.50) | 0.09<br>(0.91) | LysoPC mainly SFA and MUFA (sn1) |
| ME8 | 11 | -0.89<br>(0.27) | -1.38<br>(0.20) | -0.82<br>(0.51) | LysoPC mainly MUFA and PUFA (sn2) |
| ME9 | 25 | 0.83<br>(0.27) | -0.64<br>(0.54) | 1.45<br>(0.22) | Most abundant PCs |
| ME10 | 41 | 1.13<br>(0.18) | 1.25<br>(0.19) | 0.38<br>(0.74) | Most common cholesterol esters and sphingomyelins |
| ME11 | 27 | -0.47<br>(0.63) | -1.29<br>(0.20) | -1.18<br>(0.33) | All small TGs and in-source fragments and isotopes. |

\*upper numbers are partial coefficients extracted from the fixed effects panel model, lower numbers in parenthesis are FDR-adjusted p-values. Boxes are coloured blue when a significantly positive association was found and red when negative. Fixed effects panel models estimated the following equation $Y_{it} = \alpha_i + \beta T_i + E_{it} + \epsilon_{it}$ , with $Y_{it}$ the respective growth parameter (WAZ, LAZ, or WLZ), $\alpha_i$ the individual fixed-effect representing unobserved time-constant characteristics of the child, and the time-trend variable, which takes values between 1 (12 weeks) and 5 (104 weeks), and $E_{it}$ the respective eigenlipid.
<sup>§</sup>number of lipids belonging to the module
<sup>@</sup>PC: phosphatidylcholine, PS: phosphatidylserine, PE: phosphatidylethanolamine, TG: triglycerides, DG: diglycerides, FA: fatty acid, SFA: saturated FA, PUFA: polyunsaturated FA, MUFA: monounsaturated FA, PA: phosphatidic acid

The weighted correlation network analysis clustered lipids with very similar chemical or biological characteristics into different modules (Table 2). Most notably, triglycerides (TG) with polyunsaturated (n > 5) fatty acid (PUFA) side chains (ME6) were clustered differently from shorter-chain TGs (ME11) and TG with PUFA containing fewer double bonds (n < 4). The most abundant phosphatidylcholines (PC) found in serum were clustered in ME9, whereas cholesterol esters and sphingomyelins were clustered in ME10. Cholesterol esters and PCs with PUFA side chains however were clustered in a different module (ME2). LysoPC with saturated FA (ME7) were also clustered differently from lysoPCs with PUFA (ME8). Oxidised PCs and ether-linked PCs were clustered in ME1. Finally, any lipid that did not belong to any other module was clustered in ME5. However, these lipids also shared common characteristics such that this module is composed of free FAs and FA oxidation products and their esters. Therefore, this module cannot be discounted. Each module had a characteristic progression from 12 to 104 weeks of infant age, where the biggest changes occurred between week 12 and week 24 (Figure 2e).

#### Lipids associated with LAZ

Adjusting for age, we did not find any MEq significantly associated with LAZ over time. However, associations did appear when age in weeks (included in the model as a time trend) was removed. This indicates that the eigenlipids were significantly associated with other factors changing through time but not with LAZ itself.

We also did not find any significant differences in the MEq progression through time among the three LAZ clusters, indicating that general lipid progression is similar in all the children in this population through the first 2 years of life (Figure 3a). Multidimensional scaling analysis shows that MEq induced a time-dependent clustering of the observations, but no latent class-specific clustering is evident (Figure 3b). Analysis of individual lipid species instead of MEq also showed that lipid progression was not dependent on the LAZ growth trajectory (Supplementary Table 3).

**Figure 3.**
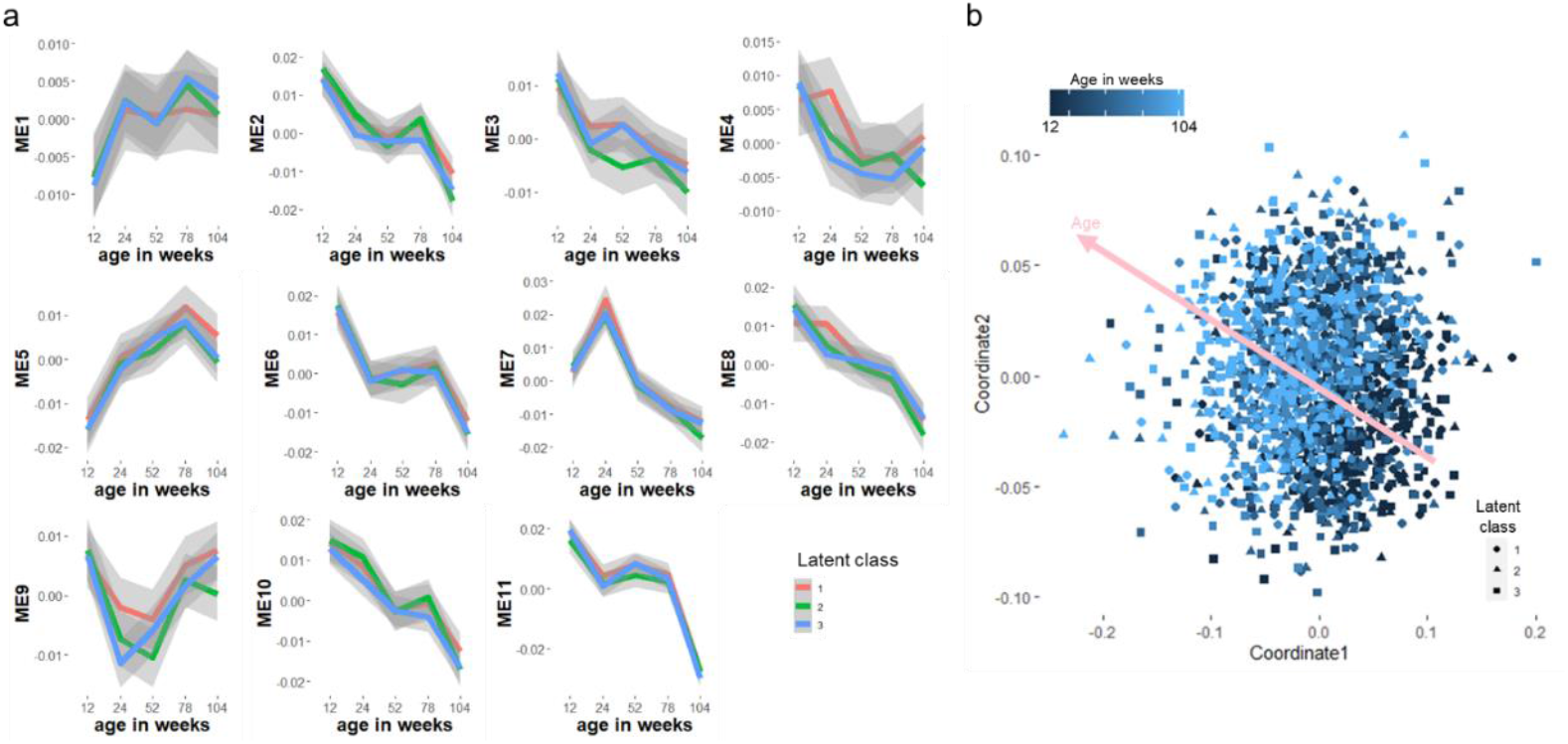
Eigenlipid progression of the children grouped based on latent class linear mixed modelling. (a) Each facet represents a module obtained from weighted correlation network analysis. No significant differences in the time-course progression of MEq were observed among the 3 clusters in all modules. (b) Multidimensional scaling analysis showing time-dependent clustering of observations but no distinction between latent classes

#### Lipids associated with WAZ and WLZ

WAZ and WLZ were highly correlated in all time points (r = 0.83, p < 0.001). Consequently, similar modules are associated with these anthropometric measures. Adjusting for age, serum levels of oxidized and ether-linked PCs (ME1) and free fatty acids (ME5) tend to have an opposite trend with WAZ and WLZ progression (p < 0.001). Conversely, the progression of PUFA-containing lipids (ME2) tended to have the same trends as WAZ and WLZ (p < 0.001) over time. As highlighted, almost all children followed the same WAZ and WLZ growth trajectory, except for a very small number of children who have improved growth parameters over time. Hence, we did not compare differences in lipid progression between these WAZ and WLZ latent classes, as there would not have been enough observations in the first cluster to make a reliable comparison.

### Panel vector autoregressive (PVAR) model using system GMM approach

We performed dynamic panel data analysis using panel vector autoregressive modelling to investigate a potential causal relationship between lipids and growth outcomes. A first order PVAR model (lag t-1) was selected as optimal lag length based on the model selection procedure of Andrews and Lu (2001)^17^. Supplementary Table 4 explores the potential causal relationship based on system GMM-PVAR model for the 3 growth outcomes and 11 lipid modules. We visually represented the results as a temporal network as shown in Figure 4. In this temporal network, current (Y_t_) and lagged values (*Y*_t-1_) of growth outcomes and each lipid module are combined into individual nodes, which are connected with directed edges according to the regression parameters (coef) of the model (Supplementary Table 4). The direction of the arrows indicate that current values of a node is consistently associated to the next (t+1) value of the other node, or itself in case of a loop. Full arrows indicate a positive association while dashed arrows indicate a negative association. Only significant (p < 0.05) associations are shown, which mean that the observed association is consistent for every time point. Hansen test for over-identifying restrictions did not reject the null hypothesis, implying that all instruments used are valid. The stability of the PVAR was confirmed as the eigenvalues as strictly less than 1 and none of the roots are outside the unit circle (Supplementary Figure 1), indicating that the model is stable and also that our variables are stationary^24,25^.

**Figure 3.**
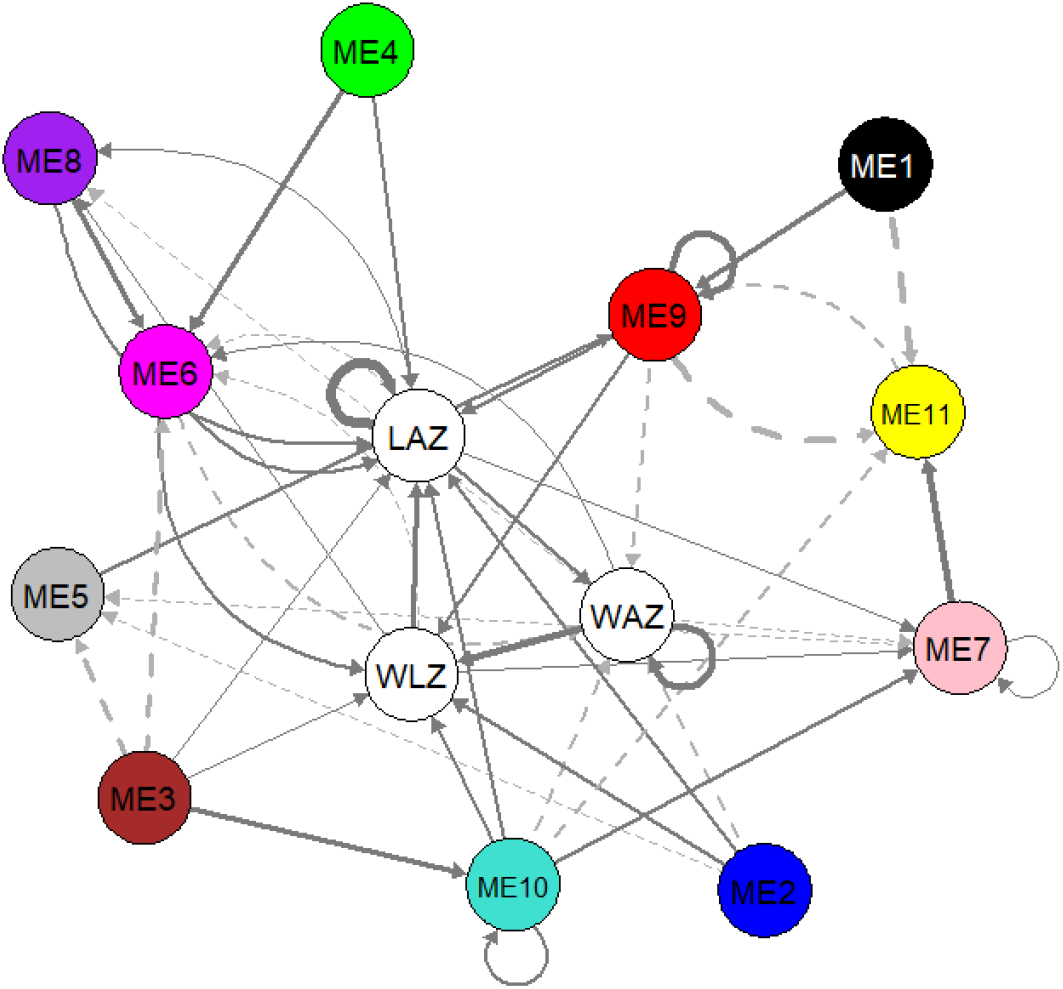
Results of system GMM-PVAR analysis. Temporal network visualization of the system GMM-based panel vector auto-regression model. Arrows indicate that a node predicts another node (or itself) in the next time point. Full arrows indicate positive while dashed arrows indicate negative association. Loops indicate that current value of a node predicts the future value of itself. Arrow thickness depicts the strength of the association. WAZ = weight-for-age z-score, WLZ = weight-for-length z-score, LAZ = length-for-age z-score, node annotation for ME1 – ME11 are shown in Table 2. All roots are inside the unit circle indicating stability of the model and stationarity of the variables (Supplementary Figure 1). WAZ, LAZ, WLZ and all ME eigenlipids were included in the model as endogenous variables in the first order (lag −1). Sex variation was accounted by adjusting for sex as an exogenous variable.

Further, the results indicate that the growth parameters positively influence each other to varying degrees; gains in WAZ increases future WLZ and WAZ itself, whereas gains in WLZ will most likely result in an increase in LAZ. Length growth also positively influences gains in weight. Modules comprising of PUFA-rich lipids (ME2 and ME6), phosphatidylcholines (ME2 and ME9), triglycerides (ME2 and ME6) and cholesterol esters (ME2 and ME10) positively impacted WLZ. However, these same lipids negatively impacted WAZ in a population with high burden of growth faltering.

Most lipid modules had positive causal links with LAZ, indicating that more biological processes and building blocks are demanded to increase height rather than weight. In addition to the lipids influencing WLZ, LAZ is also influenced by serum levels of phosphatidic acid and phosphatidylethanolamine (ME4) and lysoPC containing MUFA and PUFA lipids (ME8). ME9 had the biggest positive contribution to LAZ and WLZ among all the lipids. These positive causal relationships indicate that overall changes in the serum levels of these lipids will likely induce a change in LAZ and WLZ in the same direction; increasing serum levels of these lipid groups may lead to an increase in LAZ and WLZ.

Ether-linked phosphatidylcholines (ME1), free fatty acids (ME5), SF/MUFA-lysoPCs (ME7) and small triglycerides (ME11) did not show significant influence on any of the growth parameters. However, these lipids were shown to influence the levels of the other lipids. For instance, levels of ME1 and ME5 increase ME9 levels, whereas ME11 decreases ME9. Increasing ME7 led to a decrease in ME5 but an increase in ME11. The interplay of the lipids indicate the dynamic interactions, including synthesis and oxidation cycles, that occur between the lipid groups.

## Discussion

In this study, we introduced a statistical technique typically used in econometrics and social sciences to elucidate causal associations among growth outcome parameters and plasma lipids in the first 2 years of life of African children living in an area with a high burden of growth faltering. Using PVAR, we showed which lipids are influential to growth and also how different lipids influence each other over time.

We first characterized the children’s growth patterns in clusters using latent class mixed modelling. Previous studies that have compared growth parameters in children with metabolites have typically characterized children as either growth impaired (e.g. stunted or underweight) or healthy, and determined which metabolites or lipids are able to classify them based on this binary classification^7,26^, even for studies that observed children over a period of time^14^. Using latent class modelling, we showed that the children from rural Gambia experienced a general decline in growth outcome over time, albeit at different trajectories. This observed growth faltering has been reported previously for this cohort of children^27,28^ and is also observed to be common among children in low and middle-income countries^29^. However, while some children remained stunted over the first 2 years of life, some children remained above the stunting cut-off (despite reduced growth) while others started as normal and slowly faltered ending up as stunted in the long term. This indicates that a binary classification (impaired vs healthy) for growth faltering does not adequately capture the growth trajectories in these children.

For weight measures (WLZ and WAZ), we observed a general decline, except in a very small number of children who increased in WAZ/WLZ over time, which makes statistical comparison difficult. Future studies to investigate the progression of serum lipids among those with different WAZ/WLZ trajectories therefore require a much bigger sample size to capture enough number of children in both groups.

In Supplementary Table 5, we show that a number of studies have followed the metabolic status of children through the early years of life, especially in the first 2 years. However, most of these studies focused on well-nourished populations. A notable exception to this was the study by Giallourou et al^14^ that followed the changes that occur in the metabolome of children by analysing urine and plasma samples at 3, 6, 9, 15 and 24 months of age among children in 3 resource-constrained countries (Peru, Bangladesh and Tanzania) ^14^. The authors used a phenome-for-age z-score (PAZ) and found that PAZ of stunted children lagged compared to healthy children indicating poor metabolic maturity. These studies mainly used linear mixed/multi-levels models, ANOVA and other multidimensional data analysis techniques (PCA, partial least squares regression, ASCA). Although associations with growth outcomes can be deduced using these methods, they typically do not show variable interrelatedness and do not assess potential causal links between the metabolome/lipidome and growth outcomes.

Similar to Nikkila et al^15^, who studied serum lipidome progression among Finnish children from birth to 2 years, we began by clustering tightly correlated lipids into modules to reduce dimensionality. To do this, we used weighted gene correlation network analysis^22^. We observed that the algorithm clustered lipid species based on their chemical features, specifically type of lipid species, length and (un)saturation (SFA, MUFA, PUFA). These clusters indicate that serum levels of these lipids behave very similarly across time in the population enabling us to generalize their association with growth outcomes. We observed that major metabolic changes occurred around the first 6 months of life, which corresponded to the start of the transitional feeding in our study population – when children started taking other foods apart from breastmilk. In this population of infants, rates of exclusive breast feeding (EBF) are high, with a mean duration of EBF across the whole of the ENID cohort of 5.2 months ^27^.

Here, we treated longitudinal biochemical data as panel data, a typical data analysis concept in econometrics. Panel data is a hybrid of cross-sectional and time-series analysis, where data is collected for N individuals over T occasions, which is typically the most common design used in longitudinal systems biology studies. Systems biology can benefit from panel data analysis as is it best suited for studies with large N but small T, which is most common especially in clinical studies where participants are not able to provide numerous biological samples over long periods of time. To the best of our knowledge, this paper is the first to demonstrate the use of panel data analysis tools in systems biology.

Despite observing three categories of LAZ trajectories, we found no significant differences in the serum lipid profiles of these children over time, which concurs with our basic panel analysis (Table 2). Contrary to LAZ, we observed that lipolysis products (free fatty acids, oxidised PCs) progress reciprocally to the progression of WAZ and WLZ. However, this association does not imply that free fatty acids and oxidised PCs cause the decline in WAZ and WLZ in the first 2 years of life. One requirement for establishing a causal relationship is to demonstrate consistent significant association between current levels of the exposure and future levels of the outcome. One main advantage of using dynamic panel analysis strategies is its ability to establish potential causal links between outcomes and variables in a longitudinal study^30^. This is achieved by incorporating lagged (t-1…n) values as independent variables in the model, which will lead to biased estimates when performed using ordinary linear regression models due to the Nickell bias^20^. In econometrics, this bias is eliminated through using generalized method of moments (GMM) in dynamic panel analysis ^16,18,19,31^.

In this study, we employed a system GMM-based panel vector autoregression (PVAR) model, which allowed us to simultaneously assess the potential causal links between serum lipid profile and growth outcomes, and also how different lipid species influence each other over time. PVAR is a modification of the conventional VAR model, which deals with panel data that typically comprise designs with N > T^32^. PVAR also addresses individual heterogeneity from each individual cross-sectional unit (in this case, each child)^32^. Hence, using this method, we are able to establish potential causal links and also assess variable interrelatedness, which previous longitudinal studies in children fail to report.

Our results suggest a potential causal association between being underweight to being wasted and subsequently stunted. In a compilation of datasets from 1.8 million children in 51 countries, it was previously reported that all children that were both stunted and wasted were also underweight^33^, indicating a cross-sectional association among the 3 growth parameters. However, it has also been previously demonstrated that wasting precedes stunting and children with low WLZ were at a higher risk of linear growth retardation (stunting) especially for those below three years old^34-36^. Wasting at younger age (from 6 – 17 months) was associated with stunting from 18 months of age. This association was however not observed when wasting occurred below 6 months of age^34,35^. These earlier reports indicate that the association between the three growth outcomes was accurately captured by the PVAR model, indicating the validity of our approach.

In the interpretation of the lipid data, it is important to understand that circulating lipids in the first years of life play a crucial role growth and develop of many vital organs, most of all the brain, which requires lipid for growth and myelination. However, most information we have in the literature is still mainly limited to European or other high income studies. In a European study, full term infants fed a higher level (3.2%) of alpha-linolenic acid (ALA) during the first 4 months of life had higher plasma levels of docosahexaenoic acid (DHA) and lower mean group weight than infants on a 0.4% ALA formula. These results concur with our PVAR model indicating a negative causal link between PUFA-rich lipids (ME2 and ME6) and WAZ.

Our results demonstrated that in this population, the majority of lipids contributed to LAZ, indicating higher energy and biochemical requirements for increasing linear growth than increasing weight. In fact, WLZ alone is insufficient to influence future LAZ and several lipid clusters are needed to improve LAZ. This shows that more factors are associated with stunting than is explained by prior wasting, as also previously hypothesized^34^. The different classes of lipids involved indicate that it is not only the lipids that provide energy that are limiting growth. Most notably, lysoPCs comprised of MUFA and PUFA (ME8) were exclusively positively causal to LAZ compared to WLZ and WAZ. Evidence on the effect of PUFA, especially DHA, prenatal supplementation on infant height has been inconsistent^37^. In one study, prenatal DHA supplementation resulted to a significant increase in infant height at age 18 months compared to placebo^38^ but this effect was no longer observed when the children were followed to 60 months of age^39^. Moreover, cord blood PUFA levels were found to have a sex-specific association with infant height at 6 month of age, where n-3 PUFA levels were associated with higher infant length in males while higher n-6 PUFA concentrations were associated with lower length in infancy. However, higher cord blood n-3:n-6 ratio was associated with higher infant length at 6 months of age. These associations were however no longer observed at later time points (from 2 years of age)^40^. It is important to interpret these results in relation to nutrient availability. Brain development and growth requires large amounts of PUFAs as the brain’s lipid composition comprises 35% PUFAs, which cannot be synthesized *de novo*^41^. Hence, insufficient PUFA intake may require the body to use energy for fatty acid desaturation to enable brain development and growth, limiting the energy available for lateral growth. Supplementation with PUFAs can therefore have very different effects on growth depending on the availability of other nutrients. Hence, the potential effect of PUFA on LAZ may not be consistent. For instance, we have previously shown that PUFA supplementation did not improve growth and cognitive function of breast-fed infants in The Gambia despite increasing plasma PUFA levels^42^. PUFA intake was therefore not the limiting factor.

Of all lipids contributing to LAZ, phosphatidylcholines (ME9) has the highest influence on LAZ and WLZ. A metabolomics study reported reduced urinary levels of betaine and dimethylglycine, which are endogenous choline metabolites, in stunted Brazilian children, indicating possible reduction of choline bioavailability from the diet^26^. Choline is an essential nutrient and is a precursor for phosphatidylcholines. Low serum choline was also previously reported to be associated with linear growth failure among children in Malawi^43^. Eggs, particularly the egg yolk, are one of the main sources of dietary choline^44^. Clinical trials using egg supplementation reported improved LAZ and height gain among children in Ecuador^45^ and Uganda^46^, respectively. Although eggs contain many other important nutrients, our data suggests that this efficacy could be due at least in part to the increase in intake of phosphatidylcholine precursors.

Our current results demonstrate that all lipids species containing PUFAs (ME2, ME6 and ME8) and phosphatidylcholines (ME2, ME9) were positively contributing to infant LAZ in the first 2 years of life. This underlines the importance of availability of essential lipids in early life nutrition in these populations. More importantly, this highlights the need to use evidence from studies in the target populations, rather than relying on evidence of just European studies. Growth faltering among children in LMICs occur at a population-level^29^, which indicates the need to study its determinants at a community-level instead of looking at individuals. As the majority of these children in our study were exclusively breastfed until 5 months of age, poor maternal breastmilk lipid composition could be an underlying factor associated with growth faltering. A survey of breastmilk composition from mothers in area with high burden of infant growth faltering is therefore warranted, and could be a target for intervention.

Furthermore, environmental factors potentially contribute to the malabsorption of PUFAs and choline in these children. Environmental enteric dysfunction is a subclinical state of intestinal inflammation commonly observed in children in LMICs^47^, which may affect absorption of these lipids from breastmilk. Hence, efforts to improve sanitation and reduce incidence of infections in children may improve bioavailability of essential lipids, which leads to improved growth outcomes.

Although this paper greatly contributes to the very limited data available on the interaction between lipids and growth outcomes in the first 2 years of life, we acknowledge that our study would be improved if children with more variable growth trajectories were included. In this study population, most children exhibited very similar growth trajectories and most children were growth impaired, especially stunted. Future studies involving children with different growth outcomes within the same population is therefore warranted.

In this study, we used a high-throughput lipidomics method, which does not provide a more thorough lipid identification compared to liquid chromatography-mass spectrometry-based techniques. However, the weighted correlation network analysis allowed us to cluster lipids with similar structural and biochemical properties, which compensates for the lack of specificity in individual lipid identifications.

The causal link between underweight and wasting to stunting indicates that measures and interventions to address childhood stunting may require prevention of underweight and wasting earlier in life. Demonstrating the role of circulating lipids in growth regulation among infants in low-resource areas offers insights into potential intervention strategies based on nutritional formulation with specific lipid compositions or those that trigger increase in circulating levels of specific lipid species, especially PUFAs and PCs.

## Methods

### Study Population

The analyses presented included data and samples collected as part of the Early Nutrition and Immune Development (ENID) study, a randomised trial conducted in the rural West Kiang region of The Gambia between April 2010 and February 2015. The full ENID trial protocol is described in Moore et al ^21^ and the trial was registered as ISRCTN49285450. Briefly, mother-infant pairs were recruited in pregnancy (< 20 weeks gestation) and followed until two years post-partum. During pregnancy, women were randomly assigned to four trial arms, comparing combinations of protein-energy and multiple micronutrients and from six to 18 months of age, their infants received either a daily multiple micronutrient enriched lipid based nutritional supplement (LNS) or a placebo LNS. As part of the trial design, infant anthropometry and blood samples were collected at clinic visits at 12, 24, 52, 78 and 104 weeks of infant age. Full details of measurement and sample collection protocols can be found in the trial protocol^27^. The analyses presented here were not planned in the original study design and used data and samples from the first 400 infants born into the ENID trial.

The ENID trial was approved by the joint Gambian Government / MRC Unit The Gambia Ethics Committee (projects SCC1126v2 and L2010.77) and written, informed consent was obtained from all the participants prior to enrolment.

### Untargeted lipidomics analysis

Serum samples were stored at −80°C until assay. Lipids were extracted as described previously^48^. Briefly, 100 µl of LC–MS grade water and 150 µl of internal standard mix were added to 15 µl of serum in a 96 well glass coated plate prior to mixing for 10 s. Subsequently, 750 µl of LC–MS grade methyl-tertiary butyl ether (MTBE) and a further 200 µl of LC–MS grade water were added to each well before shaking for 10 s. Once mixed, plates were spun at 845×g for 2 min to achieve phase separation with 25 µl of the upper organic phase transferred to a new glass coated plate with 90 µl of MS-mix (7.5 mM ammonium acetate in IPA:CH3OH 2:1), which was subsequently added to each well.

### DIMS lipidomic profiling

Samples were infused into an Exactive Orbitrap (Thermo, Hemel Hempstead, UK) using a Triversa Nanomate (Advion, Ithaca, USA). Data collection began 20 s after the infusion began, initially analysing samples in the positive ionisation mode with an ionisation voltage of 1.2 kV applied. Data was acquired between 150 and 2000 m*/z* with a scan rate of 1 Hz giving a mass resolution of 65,000 at 400 m*/z*. A more detailed description of the instrument parameters can be found in Harshfield, et al. ^48^.

### Processing lipidomic data

Raw data files were converted to .mzXML files using msConvert (ProteoWizard)^49^, and were subsequently processed in R (version 3.2.2) using an in-house script to compare spectra against a list of 1649 lipid species, with a relative intensity and mass deviation value recorded for each lipid in every sample. We applied 4 filtering steps for quality control of the data and focus subsequent analysis on analytically robust signals. The first step was to remove lipids with a mean mass deviation between expected and recorded mass of greater than 5 ppm, the second step was to remove signals with an average intensity in the samples less than 5 times greater than in the blanks. The third step was to remove signals with 0 values in greater than 10% of samples. The final step was to remove lipids with an r < 0.9 in our QC dilution series.

### Data analysis

#### Analysis of growth outcomes

The changes in WAZ, LAZ and WLZ over time were determined using fixed effect panel model using growth outcomes as:

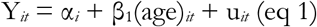

Where α_*i*_ (*i* = 1…n) is the intercept for each child, Y_*it*_ is either WAZ, LAZ or WLZ for a given child (*i*) at a particular age (*t*), and u_*it*_ is the error term. This was implemented using the plm package^50^ in R (version 3.6).

We then clustered the children based on the growth patterns using latent class mixed modelling implemented using the lcmm^51^ package. LAZ, WAZ and WLZ values of the children in all the time points were used as dependent variables, while sex and age were independent variables. Missing measurements were considered missing-at-random and hence children with incomplete measurements were included. Age and child ID were used as random effect to allow varying intercepts and slopes per individual time series. A 5-quantile splines function was employed for estimation. The number of latent classes was tested between 2 to 4 and model selection was based on the Akaike information criterion (AIC). For estimating LAZ, a 3 latent class-model yielded the least AIC value whereas a 2 latent class-models yielded least AIC values for both WAZ and WLZ.

#### Correlation network analysis

To reduce data complexity, clusters of tightly correlated lipids were determined using weighted co-expression network analysis (WGCNA^22^). Scale-free topography typical of biological networks (r^2^ ≳0.8)^23^ for our data was achieved using β = 18 for a signed network. A Pearson correlation (s_*ij*_) matrix was then generated between each lipid pairs (*i* and *j*), which was transformed into an adjacency matrix through the power transformation,

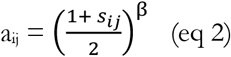

This power transformation punishes weak and negative correlations while amplifying strong positive correlations. As this study aimed to determine the dynamic changes in lipids over time, a signed network was used to determine lipids that move in the same direction over time. Using hierarchical clustering embedded with the WGCNA package, tightly correlated lipids are clustered into modules using the *blockwiseModules* function, setting the minimum number of lipids forming a module to 10. The network was visualized using igraph^52^.

#### Association between modules and growth outcomes

Each member of the module is characterized by an eigenlipid (E^(q)^, where (q) denotes the module) through a singular value decomposition. The E^(q)^ represents the collective behaviour of the particular module^23^. The progression of individual lipids or module E^(q)^ in the first 2 years of life was assessed using fixed effects panel model as in eq 1 but with E^(q)^ as additional independent variable:

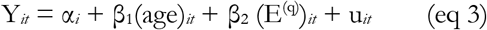

Where (E^(q)^)_*it*_ indicates the E^(q)^ of child *i* at time *t*. Significant associations of growth outcomes and lipids through time were detected using p < 0.05 after adjusting for false discovery rate (FDR) ^53^.

### Panel vector autoregression model

A PVAR model uses lags of endogenous variables and analyses interdependencies among variables of interest (LAZ, WAZ, WLZ and 11 lipid modules obtained from the weighted correlation network analysis). We thus estimated a 14-variate PVAR model of order p with panel-specific fixed effects represented by the following equation:

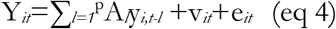

where Y_*it*_ is a (1 ×14) vector of endogenous variables for the *i*th cross-sectional unit (child) at time *t*; y_*i,t-l*_ be an 14×1 vector of lagged endogenous variables (*l* being number of lags); v_*it*_ and e_*it*_ are (1 × 14) vectors of dependent variable-specific fixed-effects and idiosyncratic errors, respectively. A_l_ represents the 14×14 matrix of endogenous parameters to be estimated.

Following the procedure of Sigmund and Ferstl (2019)^25^, we used unbalanced panel data and estimated PVAR models by fitting a multivariate panel regression of each dependent variable on lags of itself using generalized method of moments (GMM). GMM specification requires stationarity which means that all unit roots of the PVAR model fall should inside the unit circle.

The PVAR model was specified by first specifying the maximum lag order of the model using the method described by Andrews and Lu (2001)^17^. Due to maximum *t* = 5, we only tested for either first order (*t*-1) and second order (*t*-2) panels. Lag selection was based on the AIC and BIC criteria. Then, a first difference and system-GMM approaches with either first different or forward orthogonal deviation (fod) transformation were assessed. The stability of the model was then tested. The system-GMM model with fod transformation yielded a stable model and was hence used in the final analysis. The PVAR model was generated using the package *panelvar* in R^25^.

## Supporting information

Supplementary files_compiled

## Data Availability

The raw data will be published together with the manuscript. Raw data can be requested from the corresponding author.

## Acknowledgement

We thank the women of West Kiang who patiently participated in the ENID Trial. We acknowledge the enthusiastic work of the ENID study team, especially the fieldworkers, village assistants, midwives, clinical staff, data office staff and laboratory technicians who tirelessly collected the data and samples.

