## Supplementary files_compiled for "Plasma lipids and growth faltering: a longitudinal cohort study in rural Gambian children"

Supplementary figure 1. Unit root circle for the PVAR-system GMM model

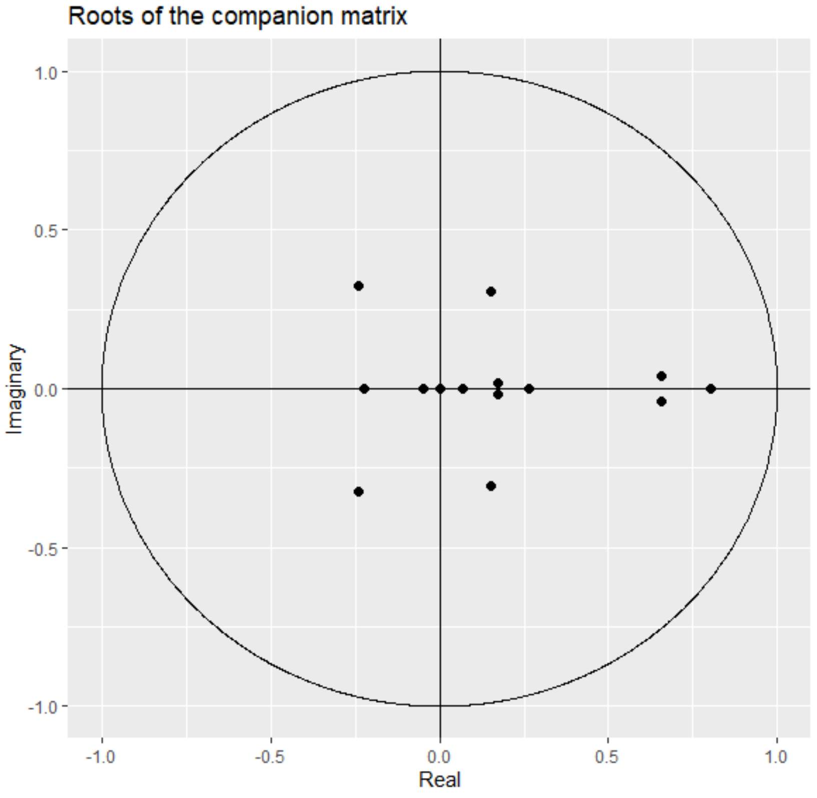

Supplementary Table 1. Individual lipid trend over the time course

**SIGNIFICANT INCREASE OVER TIME**

| ESI mode | Lipid | m/z | estimate | pVal | FDRpVal | bonferonni |
|---|---|---|---|---|---|---|
| NEG | FA(18:1)-OH_[M-H]1- | 297.2435 | 0.012 | 1.90E-71 | 2.64E-70 | 5.27E-69 |
| NEG | PE_42:5_[M-H]1- | 820.5862 | 0.012 | 1.01E-61 | 1.04E-60 | 2.82E-59 |
| NEG | OCT(C18H34O4)_[M-H]1- | 313.2384 | 0.011 | 1.15E-51 | 8.39E-51 | 3.19E-49 |
| POS | PC-O_34:3_[M+H]1+ / PE-P_37:2_[M+H]1+ | 742.5745 | 0.011 | 5.39E-57 | 4.83E-56 | 1.50E-54 |
| POS | PG_44:6_[M+H]1+ | 879.611 | 0.010 | 3.56E-41 | 1.68E-40 | 9.90E-39 |
| POS | PC_34:3_[M+H]1+ / PE_37:3_[M+H]1+ / PA_39:4_[M+NH4]1+ | 756.5538 | 0.009 | 8.10E-38 | 3.47E-37 | 2.25E-35 |
| POS | PC_34:1_[M+H]1+ / PE_37:1_[M+H]1+ / PA_39:2_[M+NH4]1+ | 760.5851 | 0.008 | 6.26E-33 | 2.20E-32 | 1.74E-30 |
| NEG | OCT(C18H32O4)_[M-H]1- | 311.2228 | 0.008 | 1.86E-27 | 5.75E-27 | 5.17E-25 |
| POS | PC_33:1_[M+H]1+ / PE_36:1_[M+H]1+ / PA_38:2_[M+NH4]1+ | 746.5694 | 0.008 | 1.02E-27 | 3.22E-27 | 2.84E-25 |
| NEG | PC_37:3_[M+Cl]1- | 832.5629 | 0.007 | 2.76E-25 | 7.78E-25 | 7.68E-23 |
| NEG | PG-P_42:6_[M-H]1- | 833.5702 | 0.007 | 9.48E-25 | 2.61E-24 | 2.64E-22 |
| POS | PI-P_30:1_[M+NH4]1+ | 782.5178 | 0.007 | 1.02E-24 | 2.77E-24 | 2.83E-22 |
| NEG | PC-P_35:2_[M+Cl]1- | 790.5523 | 0.007 | 8.66E-26 | 2.51E-25 | 2.41E-23 |
| NEG | PC_34:1_[M+OAC]1- / PS_38:0_[M-H]1- | 818.5917 | 0.007 | 1.38E-24 | 3.72E-24 | 3.83E-22 |
| NEG | SM_33:2_[M+OAC]1- | 745.5501 | 0.007 | 3.27E-20 | 7.69E-20 | 9.08E-18 |
| POS | PC-O_36:2_[M+H]1+ / PE-P_39:1_[M+H]1+ | 772.6215 | 0.006 | 1.05E-21 | 2.61E-21 | 2.92E-19 |
| POS | PC-O_34:2_[M+H]1+ / PE-O_37:2_[M+H]1+ | 744.5902 | 0.006 | 1.12E-17 | 2.36E-17 | 3.12E-15 |
| POS | PC_35:1_[M+H]1+ / PE_38:1_[M+H]1+ / PA_40:2_[M+NH4]1+ | 774.6007 | 0.006 | 2.29E-17 | 4.78E-17 | 6.36E-15 |
| NEG | FA(18:2)-OH_[M-H]1- | 295.2279 | 0.006 | 3.45E-16 | 6.91E-16 | 9.60E-14 |
| NEG | FA(18:1)_[M-H]1- | 281.2486 | 0.006 | 2.00E-15 | 3.91E-15 | 5.55E-13 |
| NEG | PC_41:6_[M+Cl]1- | 882.5785 | 0.006 | 2.37E-15 | 4.53E-15 | 6.58E-13 |
| POS | PS-O_36:2_[M+H]1+ / PG-O_36:4_[M+NH4]1+ | 774.5643 | 0.005 | 2.10E-14 | 3.92E-14 | 5.84E-12 |
| NEG | PC_39:4_[M+Cl]1- | 858.5785 | 0.005 | 1.57E-14 | 2.94E-14 | 4.36E-12 |
| POS | PC_32:1_[M+H]1+ / PE_35:1_[M+H]1+ / PA_37:2_[M+NH4]1+ | 732.5538 | 0.005 | 3.42E-14 | 6.33E-14 | 9.49E-12 |
| NEG | FA(20:1)_[M-H]1- | 309.2799 | 0.005 | 4.00E-13 | 7.12E-13 | 1.11E-10 |
| NEG | PC-O_36:3_[M+Cl]1- | 804.5679 | 0.005 | 6.34E-11 | 1.04E-10 | 1.76E-08 |
| NEG | PC-O_40:4_[M+Cl]1- | 858.6149 | 0.004 | 5.67E-10 | 9.01E-10 | 1.58E-07 |
| POS | SM_42:1_[M+H]1+ | 815.7001 | 0.004 | 2.27E-10 | 3.65E-10 | 6.31E-08 |
| NEG | PA_44:5_[M-H]1- | 805.5753 | 0.004 | 9.05E-10 | 1.43E-09 | 2.51E-07 |
| NEG | FA(16:0)_[M-H]1- | 255.233 | 0.004 | 4.54E-09 | 7.09E-09 | 1.26E-06 |
| NEG | PC_39:3_[M+Cl]1- | 860.5942 | 0.004 | 9.05E-09 | 1.38E-08 | 2.52E-06 |
| NEG | FA(16:1)_[M-H]1- | 253.2173 | 0.004 | 1.19E-08 | 1.80E-08 | 3.32E-06 |
| NEG | PC_34:2_[M+OAC]1- / PS_38:1_[M-H]1- | 816.576 | 0.004 | 3.73E-08 | 5.51E-08 | 1.04E-05 |
| POS | PC_31:2_[M+H]1+ / PE_34:2_[M+H]1+ / PA_36:3_[M+NH4]1+ | 716.5225 | 0.004 | 1.20E-07 | 1.73E-07 | 3.34E-05 |
| NEG | PC-O_36:2_[M+Cl]1- | 806.5836 | 0.004 | 5.16E-08 | 7.50E-08 | 1.43E-05 |
| POS | Cer_42:1_[M+H]1+ | 650.6446 | 0.004 | 4.71E-08 | 6.89E-08 | 1.31E-05 |
| NEG | PI_39:4_[M-H]1- | 899.5655 | 0.004 | 3.22E-07 | 4.52E-07 | 8.94E-05 |
| POS | PC-O_36:3_[M+H]1+ | 770.6058 | 0.003 | 3.45E-07 | 4.82E-07 | 9.60E-05 |
| NEG | FA(14:1)_[M-H]1- | 225.186 | 0.003 | 8.35E-07 | 1.15E-06 | 0.000232 |
| POS | CE_18:3_[M+NH4]1+ | 664.6027 | 0.003 | 3.79E-06 | 5.09E-06 | 0.00105 |
| NEG | FA(19:1)_[M-H]1- | 295.2643 | 0.003 | 3.75E-06 | 5.06E-06 | 0.00104 |
| NEG | FA(18:0)_[M-H]1- | 283.2643 | 0.003 | 6.20E-06 | 8.28E-06 | 0.00172 |
| POS | PC_34:2_[M+H]1+ / PE_37:2_[M+H]1+ / PA_39:3_[M+NH4]1+ | 758.5694 | 0.003 | 8.50E-06 | 1.13E-05 | 0.00236 |
| POS | PC_33:2_[M+H]1+ / PE_36:2_[M+H]1+ / PA_38:3_[M+NH4]1+ | 744.5538 | 0.003 | 1.34E-05 | 1.75E-05 | 0.00372 |
| POS | PI-O_38:3_[M+NH4]1+ | 892.6273 | 0.003 | 4.63E-05 | 5.93E-05 | 0.0129 |
| POS | PS-O_40:5_[M+H]1+ / PG-P_40:6_[M+NH4]1+ | 824.58 | 0.003 | 3.25E-05 | 4.20E-05 | 0.00903 |
| POS | PS-O_38:2_[M+H]1+ / PG-O_38:4_[M+NH4]1+ | 802.5956 | 0.003 | 8.88E-05 | 0.000112 | 0.0247 |
| NEG | PG-O_42:1_[M-H]1- | 845.6641 | 0.003 | 0.00018 | 0.000225 | 0.05 |

**SIGNIFICANT DECREASE OVER TIME**

| ESI mode | Lipid | m/z | estimate | pVal | FDRpVal | bonferonni |
|---|---|---|---|---|---|---|
| POS | SM_32:1_[M+H]1+ | 675.5436 | -0.017 | 6.35E-163 | 1.76E-160 | 1.76E-160 |
| NEG | PG-O_34:1_[M-H]1- | 733.5389 | -0.017 | 3.23E-158 | 4.49E-156 | 8.97E-156 |
| POS | PG_34:0_[M+H]1+x | 751.5484 | -0.016 | 2.04E-126 | 1.41E-124 | 5.66E-124 |
| POS | SM_36:1_[M+H]1+ | 731.6062 | -0.016 | 7.88E-127 | 7.30E-125 | 2.19E-124 |
| NEG | PG-O_38:1_[M-H]1- | 789.6015 | -0.016 | 3.51E-123 | 1.63E-121 | 9.76E-121 |
| POS | LPC_20:4_[M+H]1+ | 544.3398 | -0.016 | 6.36E-123 | 2.52E-121 | 1.77E-120 |

| | | | | | | |
|---|---|---|---|---|---|---|
| POS | SM_40:3_[M+H]1+ | 783.6375 | -0.016 | 1.42E-123 | 7.92E-122 | 3.96E-121 |
| POS | PE-O_38:6_[M+H]1+ | 750.5432 | -0.015 | 7.99E-117 | 2.22E-115 | 2.22E-114 |
| NEG | PG-O_40:1_[M-H]1- | 817.6328 | -0.015 | 5.09E-115 | 1.29E-113 | 1.41E-112 |
| POS | SM_38:1_[M+H]1+ | 759.6375 | -0.015 | 9.28E-120 | 3.23E-118 | 2.58E-117 |
| POS | SM_30:1_[M+H]1+ | 647.5123 | -0.015 | 5.47E-117 | 1.69E-115 | 1.52E-114 |
| NEG | SM_30:1_[M+OAC]1- | 705.5188 | -0.014 | 3.43E-98 | 7.94E-97 | 9.53E-96 |
| POS | DG_42:7_[M+H-H2O]1+ | 677.5503 | -0.014 | 4.11E-96 | 8.80E-95 | 1.14E-93 |
| NEG | PG-O_38:2_[M-H]1- | 787.5858 | -0.013 | 5.85E-91 | 1.16E-89 | 1.63E-88 |
| POS | SM_42:3_[M+H]1+ | 811.6688 | -0.013 | 2.00E-81 | 3.48E-80 | 5.57E-79 |
| POS | SM_39:1_[M+H]1+ | 773.6531 | -0.013 | 1.01E-82 | 1.86E-81 | 2.79E-80 |
| NEG | SM_32:2_[M+OAC]1- | 731.5345 | -0.013 | 1.60E-73 | 2.48E-72 | 4.46E-71 |
| POS | PC_38:4_[M+H]1+ / PE_41:4_[M+H]1+ | 810.6007 | -0.013 | 1.85E-76 | 3.02E-75 | 5.13E-74 |
| POS | SM_36:2_[M+H]1+ | 729.5905 | -0.012 | 2.19E-73 | 3.21E-72 | 6.10E-71 |
| POS | DG_30:1_[M+H-H2O]1+ | 521.4564 | -0.012 | 2.32E-65 | 3.07E-64 | 6.45E-63 |
| POS | PC-O_18:0_[M+H]1+ / LPE_21:0_[M+H]1+ | 524.3711 | -0.012 | 2.53E-62 | 2.70E-61 | 7.02E-60 |
| POS | PE-P_38:6_[M+H]1+ | 748.5276 | -0.012 | 2.65E-64 | 3.34E-63 | 7.36E-62 |
| POS | TG_48:3_[M+NH4]1+ | 818.7232 | -0.012 | 4.50E-63 | 5.44E-62 | 1.25E-60 |
| POS | TG_46:3_[M+NH4]1+ | 790.6919 | -0.012 | 2.44E-62 | 2.70E-61 | 6.79E-60 |
| POS | LPC_20:3_[M+H]1+ | 546.3554 | -0.011 | 5.26E-59 | 5.04E-58 | 1.46E-56 |
| POS | CE_20:4_[M+NH4]1+ | 690.6184 | -0.011 | 2.13E-62 | 2.47E-61 | 5.92E-60 |
| NEG | LPE_20:0_[M-H]1- | 508.3409 | -0.011 | 7.98E-57 | 6.93E-56 | 2.22E-54 |
| POS | TG_46:2_[M+NH4]1+ | 792.7076 | -0.011 | 1.47E-58 | 1.37E-57 | 4.10E-56 |
| NEG | PG-O_37:1_[M-H]1- | 775.5858 | -0.011 | 5.50E-61 | 5.46E-60 | 1.53E-58 |
| POS | SM_40:2_[M+H]1+ | 785.6531 | -0.011 | 6.70E-55 | 5.65E-54 | 1.86E-52 |
| POS | TG_48:2_[M+NH4]1+ | 820.7389 | -0.011 | 1.48E-53 | 1.21E-52 | 4.13E-51 |
| POS | TG_42:1_[M+NH4]1+ | 738.6606 | -0.011 | 3.47E-52 | 2.61E-51 | 9.66E-50 |
| NEG | PG-P_41:0_[M-H]1- | 831.6484 | -0.010 | 8.55E-53 | 6.60E-52 | 2.38E-50 |
| POS | DG_40:3_[M+NH4]1+ | 692.6188 | -0.010 | 7.77E-53 | 6.17E-52 | 2.16E-50 |
| POS | TG_42:2_[M+NH4]1+ | 736.645 | -0.010 | 1.47E-50 | 1.02E-49 | 4.08E-48 |
| POS | TG_44:1_[M+NH4]1+ | 766.6919 | -0.010 | 1.10E-48 | 7.47E-48 | 3.06E-46 |
| POS | TG_46:1_[M+NH4]1+ | 794.7232 | -0.010 | 1.78E-48 | 1.18E-47 | 4.95E-46 |
| NEG | PG-O_40:2_[M-H]1- | 815.6171 | -0.010 | 4.13E-51 | 2.94E-50 | 1.15E-48 |
| POS | TG_44:2_[M+NH4]1+ | 764.6763 | -0.010 | 1.91E-48 | 1.24E-47 | 5.32E-46 |
| POS | PC_38:3_[M+H]1+ / PE_41:3_[M+H]1+ / PA_43:4_[M+NH4]1+ | 812.6164 | -0.010 | 2.05E-47 | 1.27E-46 | 5.70E-45 |
| POS | SM_35:1_[M+H]1+ | 717.5905 | -0.010 | 2.77E-48 | 1.75E-47 | 7.71E-46 |
| NEG | PC-O_14:0_[M+OAC]1- | 526.315 | -0.010 | 3.45E-45 | 1.92E-44 | 9.60E-43 |
| POS | TG_40:1_[M+NH4]1+ | 710.6293 | -0.010 | 8.61E-45 | 4.69E-44 | 2.39E-42 |
| POS | PC-O_18:1_[M+H]1+ | 522.3554 | -0.010 | 5.58E-44 | 2.98E-43 | 1.55E-41 |
| POS | TG_58:7_[M+NH4]1+ | 950.8171 | -0.010 | 1.28E-45 | 7.43E-45 | 3.57E-43 |
| POS | LPC_18:2_[M+H]1+ | 520.3398 | -0.010 | 1.21E-45 | 7.16E-45 | 3.36E-43 |
| POS | TG_56:5_[M+NH4]1+ | 926.8171 | -0.010 | 8.27E-46 | 5.00E-45 | 2.30E-43 |
| NEG | PG-O_42:2_[M-H]1- | 843.6484 | -0.010 | 1.53E-42 | 7.74E-42 | 4.26E-40 |
| POS | TG_50:5_[M+NH4]1+ | 842.7232 | -0.010 | 3.17E-43 | 1.63E-42 | 8.82E-41 |
| NEG | LPE_20:4_[M-H]1- | 500.2783 | -0.010 | 2.27E-45 | 1.29E-44 | 6.31E-43 |
| POS | CE_16:0_[M+NH4]1+ | 642.6184 | -0.009 | 7.36E-42 | 3.65E-41 | 2.05E-39 |
| POS | TG_50:4_[M+NH4]1+ | 844.7389 | -0.009 | 2.22E-41 | 1.07E-40 | 6.18E-39 |
| NEG | LPE_20:2_[M-H]1- | 504.3096 | -0.009 | 1.08E-41 | 5.26E-41 | 3.00E-39 |
| POS | LPE_20:4_[M+H]1+ | 502.2928 | -0.009 | 1.04E-43 | 5.47E-43 | 2.90E-41 |
| POS | PC_40:6_[M+H]1+ / PE_43:6_[M+H]1+ | 834.6007 | -0.009 | 5.42E-39 | 2.47E-38 | 1.51E-36 |
| NEG | LPC_18:2_[M+OAC]1- / LPS_22:1_[M-H]1- | 578.3463 | -0.009 | 1.15E-38 | 5.08E-38 | 3.20E-36 |
| NEG | LPE_20:1_[M-H]1- | 506.3252 | -0.009 | 6.12E-39 | 2.74E-38 | 1.70E-36 |
| NEG | PA_41:4_[M-H]1- | 765.544 | -0.009 | 2.92E-38 | 1.27E-37 | 8.12E-36 |
| POS | TG_56:6_[M+NH4]1+ | 924.8015 | -0.009 | 3.43E-40 | 1.59E-39 | 9.54E-38 |
| POS | PE-O_36:5_[M+H]1+ | 724.5276 | -0.009 | 1.06E-36 | 4.23E-36 | 2.96E-34 |
| POS | PE-O_38:5_[M+H]1+ | 752.5589 | -0.009 | 6.17E-37 | 2.52E-36 | 1.71E-34 |
| NEG | PC-O_33:2_[M+Cl]1- | 764.5366 | -0.009 | 1.05E-36 | 4.22E-36 | 2.91E-34 |
| POS | SM_42:2_[M+H]1+ | 813.6844 | -0.009 | 3.51E-37 | 1.46E-36 | 9.76E-35 |
| POS | TG_50:3_[M+NH4]1+ | 846.7545 | -0.009 | 1.47E-37 | 6.20E-37 | 4.09E-35 |
| POS | PC-O_16:0_[M+H]1+ / LPE_19:0_[M+H]1+ | 496.3398 | -0.009 | 1.96E-34 | 7.27E-34 | 5.45E-32 |
| NEG | PC-O_16:0_[M+OAC]1- | 554.3463 | -0.009 | 2.00E-34 | 7.33E-34 | 5.57E-32 |
| POS | TG_58:8_[M+NH4]1+ | 948.8015 | -0.009 | 1.90E-36 | 7.34E-36 | 5.28E-34 |
| POS | PI_38:4_[M+NH4]1+ | 904.591 | -0.009 | 1.47E-35 | 5.51E-35 | 4.08E-33 |
| POS | DG_28:0_[M+H-H2O]1+ | 495.4408 | -0.009 | 3.88E-34 | 1.40E-33 | 1.08E-31 |

| | | | | | | |
|---|---|---|---|---|---|---|
| POS | SM_33:1_[M+H]1+ | 689.5592 | -0.009 | 5.32E-36 | 2.02E-35 | 1.48E-33 |
| POS | LPE_22:6_[M+H]1+ | 526.2928 | -0.009 | 1.56E-36 | 6.12E-36 | 4.35E-34 |
| POS | DG_34:1_[M+NH4]1+ | 612.5562 | -0.008 | 4.07E-32 | 1.40E-31 | 1.13E-29 |
| POS | PC-O_14:0_[M+H]1+ / LPE_17:0_[M+H]1+ | 468.3085 | -0.008 | 3.01E-31 | 1.02E-30 | 8.37E-29 |
| NEG | LPE_22:6_[M-H]1- | 524.2783 | -0.008 | 4.61E-33 | 1.64E-32 | 1.28E-30 |
| POS | TG_56:7_[M+NH4]1+ | 922.7858 | -0.008 | 2.01E-32 | 6.98E-32 | 5.58E-30 |
| POS | TG_56:8_[M+NH4]1+ | 920.7702 | -0.008 | 3.22E-31 | 1.08E-30 | 8.96E-29 |
| POS | TG_48:1_[M+NH4]1+ | 822.7545 | -0.008 | 2.58E-29 | 8.24E-29 | 7.17E-27 |
| POS | TG_52:6_[M+NH4]1+ | 868.7389 | -0.008 | 3.58E-30 | 1.18E-29 | 9.94E-28 |
| POS | TG_58:9_[M+NH4]1+ | 946.7858 | -0.008 | 5.40E-30 | 1.76E-29 | 1.50E-27 |
| POS | DG_26:0_[M+H-H2O]1+ | 467.4095 | -0.008 | 2.50E-27 | 7.65E-27 | 6.96E-25 |
| POS | TG_38:0_[M+NH4]1+ | 684.6137 | -0.008 | 5.66E-27 | 1.71E-26 | 1.57E-24 |
| POS | LPC_20:5_[M+H]1+ | 542.3241 | -0.008 | 1.11E-27 | 3.47E-27 | 3.09E-25 |
| POS | PE-P_40:6_[M+H]1+ | 776.5589 | -0.008 | 1.41E-26 | 4.16E-26 | 3.91E-24 |
| POS | PC-O_44:5_[M+H]1+ | 878.6997 | -0.008 | 8.29E-30 | 2.68E-29 | 2.30E-27 |
| NEG | LPE_18:0_[M-H]1- | 480.3096 | -0.007 | 9.16E-25 | 2.55E-24 | 2.55E-22 |
| POS | PC_36:4_[M+H]1+ / PE_39:4_[M+H]1+ / PA_41:5_[M+NH4]1+ | 782.5694 | -0.007 | 1.72E-26 | 5.03E-26 | 4.77E-24 |
| POS | PC_35:4_[M+H]1+ / PE_38:4_[M+H]1+ / PA_40:5_[M+NH4]1+ | 768.5538 | -0.007 | 7.86E-27 | 2.35E-26 | 2.19E-24 |
| NEG | PC-O_40:6_[M+Cl]1- | 854.5836 | -0.007 | 1.34E-25 | 3.84E-25 | 3.73E-23 |
| POS | DG_44:3_[M+H-H2O]1+ | 713.6442 | -0.007 | 3.59E-24 | 9.59E-24 | 9.97E-22 |
| POS | TG_40:0_[M+NH4]1+ | 712.645 | -0.007 | 3.79E-24 | 1.00E-23 | 1.05E-21 |
| POS | PC-O_38:5_[M+H]1+ | 794.6058 | -0.007 | 2.77E-25 | 7.78E-25 | 7.70E-23 |
| POS | SM_34:2_[M+H]1+ | 701.5592 | -0.007 | 5.72E-23 | 1.50E-22 | 1.59E-20 |
| POS | DG_30:0_[M+H-H2O]1+ | 523.4721 | -0.007 | 5.14E-22 | 1.31E-21 | 1.43E-19 |
| POS | CE_22:6_[M+NH4]1+ | 714.6184 | -0.007 | 2.74E-22 | 7.11E-22 | 7.61E-20 |
| POS | TG_56:4_[M+NH4]1+ | 928.8328 | -0.007 | 3.90E-22 | 1.00E-21 | 1.08E-19 |
| POS | MG_18:1_[M+NH4]1+ | 374.3265 | -0.007 | 8.34E-21 | 1.98E-20 | 2.32E-18 |
| POS | PC_40:7_[M+H]1+ | 832.5851 | -0.007 | 6.88E-22 | 1.72E-21 | 1.91E-19 |
| POS | PC_32:0_[M+H]1+ / PE_35:0_[M+H]1+ / PA_37:1_[M+NH4]1+ | 734.5694 | -0.007 | 3.88E-21 | 9.37E-21 | 1.08E-18 |
| POS | PC-O_36:4_[M+H]1+ | 768.5902 | -0.007 | 6.60E-22 | 1.67E-21 | 1.83E-19 |
| POS | PC_38:2_[M+H]1+ / PE_41:2_[M+H]1+ | 814.632 | -0.007 | 4.80E-21 | 1.15E-20 | 1.33E-18 |
| POS | TG_52:5_[M+NH4]1+ | 870.7545 | -0.007 | 2.74E-21 | 6.68E-21 | 7.61E-19 |
| POS | TG_54:7_[M+NH4]1+ | 894.7545 | -0.007 | 1.35E-21 | 3.31E-21 | 3.75E-19 |
| NEG | SM_36:0_[M+OAC]1- | 791.6284 | -0.007 | 1.15E-19 | 2.62E-19 | 3.20E-17 |
| POS | SM_38:2_[M+H]1+ | 757.6218 | -0.007 | 1.16E-19 | 2.63E-19 | 3.23E-17 |
| POS | TG_36:0_[M+NH4]1+ | 656.5824 | -0.006 | 2.92E-19 | 6.49E-19 | 8.11E-17 |
| NEG | LPE_16:0_[M-H]1- | 452.2783 | -0.006 | 5.16E-20 | 1.20E-19 | 1.43E-17 |
| POS | TG_54:1_[M+NH4]1+ | 906.8484 | -0.006 | 3.99E-19 | 8.79E-19 | 1.11E-16 |
| POS | SM_41:1_[M+H]1+ | 799.6688 | -0.006 | 5.66E-20 | 1.31E-19 | 1.57E-17 |
| POS | PC-O_36:5_[M+H]1+ | 766.5745 | -0.006 | 1.15E-19 | 2.62E-19 | 3.20E-17 |
| POS | TG_54:6_[M+NH4]1+ | 896.7702 | -0.006 | 2.13E-19 | 4.77E-19 | 5.92E-17 |
| NEG | PG-O_36:2_[M-H]1- | 759.5545 | -0.006 | 9.60E-19 | 2.09E-18 | 2.67E-16 |
| NEG | FA(24:1)_[M-H]1- | 365.3425 | -0.006 | 4.64E-18 | 9.85E-18 | 1.29E-15 |
| NEG | PC_36:4_[M+OAC]1- / PS_40:3_[M-H]1- | 840.576 | -0.006 | 2.86E-18 | 6.11E-18 | 7.95E-16 |
| POS | PC_37:4_[M+H]1+ / PE_40:4_[M+H]1+ / PA_42:5_[M+NH4]1+ | 796.5851 | -0.006 | 5.70E-19 | 1.25E-18 | 1.58E-16 |
| POS | DG_36:3_[M+NH4]1+ | 636.5562 | -0.006 | 1.41E-18 | 3.04E-18 | 3.93E-16 |
| POS | TG_42:0_[M+NH4]1+ | 740.6763 | -0.006 | 3.04E-17 | 6.31E-17 | 8.46E-15 |
| POS | PG_36:1_[M+H]1+ | 777.564 | -0.006 | 2.11E-16 | 4.27E-16 | 5.85E-14 |
| POS | LPC-O_18:1_[M+H]1+ | 508.3762 | -0.006 | 8.25E-16 | 1.64E-15 | 2.29E-13 |
| POS | LPE_18:2_[M+H]1+ | 478.2928 | -0.006 | 6.93E-17 | 1.42E-16 | 1.93E-14 |
| POS | TG_50:2_[M+NH4]1+ | 848.7702 | -0.006 | 5.38E-17 | 1.11E-16 | 1.50E-14 |
| POS | PG_37:0_[M+H]1+ | 793.5953 | -0.006 | 2.43E-16 | 4.90E-16 | 6.76E-14 |
| POS | TG_44:0_[M+NH4]1+ | 768.7076 | -0.006 | 2.33E-15 | 4.50E-15 | 6.48E-13 |
| POS | DG_34:2_[M+NH4]1+ | 610.5405 | -0.006 | 7.94E-15 | 1.51E-14 | 2.21E-12 |
| POS | PC-O_38:6_[M+H]1+ | 792.5902 | -0.005 | 2.16E-15 | 4.20E-15 | 6.00E-13 |
| POS | PC-O_38:4_[M+H]1+ | 796.6215 | -0.005 | 8.51E-16 | 1.68E-15 | 2.36E-13 |
| POS | TG_54:5_[M+NH4]1+ | 898.7858 | -0.005 | 5.75E-14 | 1.05E-13 | 1.60E-11 |
| NEG | PG-O_35:1_[M-H]1- | 747.5545 | -0.005 | 1.22E-14 | 2.31E-14 | 3.39E-12 |
| POS | TG_56:9_[M+NH4]1+ | 918.7545 | -0.005 | 3.78E-14 | 6.96E-14 | 1.05E-11 |
| POS | SM_34:1_[M+H]1+ | 703.5749 | -0.005 | 7.80E-14 | 1.42E-13 | 2.17E-11 |
| POS | DG_44:7_[M+H-H2O]1+ | 705.5816 | -0.005 | 2.31E-13 | 4.15E-13 | 6.43E-11 |
| POS | PC_38:6_[M+H]1+ / PE_41:6_[M+H]1+ | 806.5694 | -0.005 | 1.42E-13 | 2.56E-13 | 3.94E-11 |
| POS | PE-O_40:6_[M+H]1+ | 778.5745 | -0.005 | 1.36E-12 | 2.37E-12 | 3.78E-10 |

| | | | | | | |
|---|---|---|---|---|---|---|
| NEG | LPC_20:5_[M+OAC]1- | 600.3307 | -0.005 | 1.09E-12 | 1.91E-12 | 3.02E-10 |
| POS | SM_32:2_[M+H]1+ | 673.5279 | -0.005 | 5.17E-12 | 8.88E-12 | 1.44E-09 |
| NEG | LPC-O_16:1_[M+OAC]1- / LPS-O_20:0_[M-H]1- | 538.3514 | -0.005 | 6.92E-12 | 1.18E-11 | 1.92E-09 |
| POS | DG_42:5_[M+NH4]1+ | 716.6188 | -0.005 | 5.02E-12 | 8.67E-12 | 1.40E-09 |
| POS | PC_36:2_[M+H]1+ / PE_39:2_[M+H]1+ / PA_41:3_[M+NH4]1+ | 786.6007 | -0.005 | 1.04E-12 | 1.85E-12 | 2.90E-10 |
| NEG | SM_34:1_[M+OAC]1- | 761.5814 | -0.005 | 7.30E-12 | 1.24E-11 | 2.03E-09 |
| POS | TG_52:4_[M+NH4]1+ | 872.7702 | -0.005 | 2.66E-12 | 4.62E-12 | 7.39E-10 |
| POS | LPC-2O_16:0_[M+H]1+ | 482.3605 | -0.005 | 1.15E-10 | 1.87E-10 | 3.21E-08 |
| POS | TG_53:4_[M+NH4]1+ | 886.7858 | -0.005 | 1.32E-11 | 2.21E-11 | 3.67E-09 |
| NEG | LPE_18:2_[M-H]1- | 476.2783 | -0.005 | 6.08E-11 | 1.01E-10 | 1.69E-08 |
| POS | PC-2O_32:0_[M+H]1+ | 706.6109 | -0.005 | 3.61E-11 | 6.02E-11 | 1.00E-08 |
| POS | LPC_16:1_[M+H]1+ / LPE_19:1_[M+H]1+ | 494.3241 | -0.004 | 2.39E-10 | 3.82E-10 | 6.65E-08 |
| POS | DG_34:2_[M+H-H2O]1+ | 575.5034 | -0.004 | 7.45E-11 | 1.22E-10 | 2.07E-08 |
| POS | PC-P_42:4_[M+H]1+ | 850.6684 | -0.004 | 1.06E-11 | 1.78E-11 | 2.94E-09 |
| POS | DG_36:3_[M+H-H2O]1+ | 601.519 | -0.004 | 1.81E-10 | 2.93E-10 | 5.03E-08 |
| POS | TG_58:10_[M+NH4]1+ | 944.7702 | -0.004 | 7.15E-09 | 1.10E-08 | 1.99E-06 |
| POS | PG_39:0_[M+H]1+ | 821.6266 | -0.004 | 3.55E-09 | 5.58E-09 | 9.88E-07 |
| NEG | BA(GDCA)_[M-H]1- | 448.3068 | -0.004 | 1.14E-08 | 1.72E-08 | 3.17E-06 |
| NEG | LPC-2O_16:0_[M+OAC]1- | 540.3671 | -0.004 | 3.30E-08 | 4.90E-08 | 9.16E-06 |
| POS | PC-O_40:6_[M+H]1+ | 820.6215 | -0.004 | 4.90E-09 | 7.62E-09 | 1.36E-06 |
| POS | TG_54:4_[M+NH4]1+ | 900.8015 | -0.004 | 1.47E-08 | 2.20E-08 | 4.09E-06 |
| POS | DG_34:1_[M+H-H2O]1+ | 577.519 | -0.004 | 7.52E-09 | 1.16E-08 | 2.09E-06 |
| POS | TG_52:3_[M+NH4]1+ | 874.7858 | -0.004 | 9.01E-09 | 1.38E-08 | 2.50E-06 |
| POS | DG_36:1_[M+H-H2O]1+ | 605.5503 | -0.004 | 4.09E-08 | 6.02E-08 | 1.14E-05 |
| NEG | PC-O-31:0_[M+OAC]1- | 764.5811 | -0.004 | 8.64E-08 | 1.25E-07 | 2.40E-05 |
| NEG | FA(20:4)_[M-H]1- | 303.233 | -0.004 | 1.54E-07 | 2.20E-07 | 4.29E-05 |
| POS | Cholesterol_[M+H-H2O]1+ | 369.3516 | -0.004 | 1.31E-07 | 1.87E-07 | 3.64E-05 |
| POS | TG_49:2_[M+NH4]1+ | 834.7545 | -0.004 | 1.68E-07 | 2.39E-07 | 4.68E-05 |
| POS | TG_51:1_[M+NH4]1+ | 864.8015 | -0.004 | 4.08E-07 | 5.68E-07 | 0.000114 |
| POS | TG_54:2_[M+NH4]1+ | 904.8328 | -0.004 | 3.09E-07 | 4.35E-07 | 8.58E-05 |
| NEG | PC-O_35:2_[M+Cl]1- | 792.5679 | -0.004 | 8.49E-07 | 1.17E-06 | 0.000236 |
| POS | PC-O_32:1_[M+H]1+ / PE-O_35:1_[M+H]1+ | 718.5745 | -0.003 | 1.07E-06 | 1.47E-06 | 0.000297 |
| NEG | LPE_18:1_[M-H]1- | 478.2939 | -0.003 | 2.12E-06 | 2.88E-06 | 0.000588 |
| POS | LPC_13:0_[M+H]1+ / LPE_16:0_[M+H]1+ | 454.2928 | -0.003 | 6.80E-06 | 9.05E-06 | 0.00189 |
| POS | TG_51:3_[M+NH4]1+ | 860.7702 | -0.003 | 3.43E-06 | 4.65E-06 | 0.000953 |
| POS | PC_38:1_[M+H]1+ / PE_41:1_[M+H]1+ / PA_43:2_[M+NH4]1+ | 816.6477 | -0.003 | 1.15E-05 | 1.51E-05 | 0.0032 |
| POS | TG_53:3_[M+NH4]1+ | 888.8015 | -0.003 | 1.06E-05 | 1.39E-05 | 0.00294 |
| POS | PC-O_34:1_[M+H]1+ / PE-O_37:1_[M+H]1+ | 746.6058 | -0.003 | 3.40E-05 | 4.37E-05 | 0.00945 |
| POS | PC_40:4_[M+H]1+ / PE_43:4_[M+H]1+ | 838.632 | -0.003 | 2.56E-05 | 3.33E-05 | 0.00713 |
| POS | PC_38:5_[M+H]1+ / PE_41:5_[M+H]1+ / PA_43:6_[M+NH4]1+ | 808.5851 | -0.003 | 8.68E-05 | 0.00011 | 0.0241 |
| POS | DG_36:2_[M+NH4]1+ | 638.5718 | -0.003 | 5.45E-05 | 6.95E-05 | 0.0152 |
| POS | TG_52:2_[M+NH4]1+ | 876.8015 | -0.003 | 5.55E-05 | 7.05E-05 | 0.0154 |
| POS | TG_54:3_[M+NH4]1+ | 902.8171 | -0.003 | 0.000182 | 0.000227 | 0.0506 |

**NO SIGNIFICANT CHANGE OVER TIME**

| ESI mode | Lipid | m/z | estimate | pVal | FDRpVal | bonferonni |
|---|---|---|---|---|---|---|
| POS | CE_20:5_[M+NH4]1+ | 688.6027 | -0.003 | 0.000199 | 0.000247 | 0.0552 |
| POS | DG_36:2_[M+H-H2O]1+ | 603.5347 | -0.003 | 0.000213 | 0.000264 | 0.0593 |
| POS | PE-O_34:3_[M+H]1+ | 700.5276 | 0.003 | 0.000316 | 0.000388 | 0.0878 |
| NEG | PA_43:4_[M-H]1- | 793.5753 | -0.003 | 0.000331 | 0.000406 | 0.0921 |
| POS | PC_39:6_[M+H]1+ / PE_42:6_[M+H]1+ / PA_44:7_[M+NH4]1+ | 820.5851 | -0.003 | 0.000333 | 0.000406 | 0.0925 |
| NEG | FA(22:1)_[M-H]1- | 337.3112 | -0.003 | 0.000355 | 0.000431 | 0.0986 |
| POS | Hydroxycholesterol_[M+H-2H2O]1+ | 367.3359 | 0.003 | 0.000365 | 0.000441 | 0.101 |
| POS | PC_37:6_[M+H]1+ / PE_40:6_[M+H]1+ / PA_42:7_[M+NH4]1+ | 792.5538 | -0.002 | 0.00045 | 0.000541 | 0.125 |
| NEG | FA(22:6)_[M-H]1- | 327.233 | -0.002 | 0.001055 | 0.001264 | 0.293 |
| POS | PC_33:3_[M+H]1+ / PE_36:3_[M+H]1+ / PA_38:4_[M+NH4]1+ | 742.5381 | 0.002 | 0.001283 | 0.001531 | 0.357 |
| POS | TG_50:1_[M+NH4]1+ | 850.7858 | -0.002 | 0.00138 | 0.001639 | 0.384 |
| POS | PS_38:2_[M+H]1+ / PG_38:4_[M+NH4]1+ | 816.5749 | 0.002 | 0.001506 | 0.001781 | 0.419 |
| POS | DG_32:0_[M+H-H2O]1+ | 551.5034 | -0.002 | 0.001553 | 0.00183 | 0.432 |
| NEG | LPE-P_16:0_[M-H]1- | 436.2833 | -0.002 | 0.001639 | 0.001922 | 0.456 |
| POS | PC_36:3_[M+H]1+ / PE_39:3_[M+H]1+ / PA_41:4_[M+NH4]1+ | 784.5851 | -0.002 | 0.002134 | 0.002493 | 0.593 |
| POS | PC-P_38:6_[M+H]1+ | 790.5745 | 0.002 | 0.002476 | 0.00288 | 0.688 |

| | | | | | | |
|---|---|---|---|---|---|---|
| NEG | EIC(C20H38O4)_[M-H]1- | 341.2697 | 0.002 | 0.003355 | 0.003884 | 0.933 |
| POS | PE-O_40:5_[M+H]1+ | 780.5902 | 0.002 | 0.003367 | 0.003884 | 0.936 |
| POS | PI_40:2_[M+NH4]1+ | 936.6536 | -0.002 | 0.003727 | 0.004281 | 1.04 |
| NEG | FA(22:4)_[M-H]1- | 331.2643 | -0.002 | 0.00402 | 0.004599 | 1.12 |
| NEG | PC-O_36:4_[M+Cl]1- | 802.5523 | 0.002 | 0.004515 | 0.005144 | 1.26 |
| POS | PG_37:1_[M+H]1+ | 791.5797 | 0.002 | 0.005626 | 0.006384 | 1.56 |
| NEG | PE_44:5_[M-H]1- | 848.6175 | 0.002 | 0.006712 | 0.007585 | 1.87 |
| NEG | FAHFA_PAHPA_[M-H]1- | 509.4575 | 0.002 | 0.007892 | 0.008882 | 2.19 |
| POS | CE_18:1_[M+NH4]1+ | 668.634 | -0.002 | 0.009004 | 0.010093 | 2.5 |
| POS | TG_53:2_[M+NH4]1+ | 890.8171 | -0.002 | 0.012389 | 0.013832 | 3.44 |
| POS | PC_36:1_[M+H]1+ / PE_39:1_[M+H]1+ / PA_41:2_[M+NH4]1+ | 788.6164 | 0.002 | 0.012559 | 0.013966 | 3.49 |
| POS | DG_38:0_[M+NH4]1+ | 670.6344 | -0.002 | 0.014114 | 0.015633 | 3.92 |
| POS | PC-O_32:0_[M+H]1+ / PE-O_35:0_[M+H]1+ | 720.5902 | -0.002 | 0.015738 | 0.017362 | 4.38 |
| NEG | PC-P_36:5_[M+OAC]1- / PS-O_40:5_[M-H]1- | 822.5654 | -0.002 | 0.026215 | 0.028805 | 7.29 |
| POS | PC-O_40:4_[M+H]1+ | 824.6528 | -0.001 | 0.032401 | 0.035463 | 9.01 |
| POS | TG_51:2_[M+NH4]1+ | 862.7858 | -0.001 | 0.03662 | 0.039923 | 10.2 |
| POS | PC_34:4_[M+H]1+ / PE_37:4_[M+H]1+ / PA_39:5_[M+NH4]1+ | 754.5381 | -0.001 | 0.045565 | 0.049481 | 12.7 |
| POS | SM_41:0_[M+H]1+ | 801.6844 | 0.001 | 0.087841 | 0.095019 | 24.4 |
| POS | PC_36:6_[M+H]1+ / PE_39:6_[M+H]1+ / PA_41:7_[M+NH4]1+ | 778.5381 | 0.001 | 0.099077 | 0.106757 | 27.5 |
| NEG | PA_43:6_[M-H]1- | 789.544 | 0.001 | 0.141852 | 0.152258 | 39.4 |
| POS | Cer_42:2_[M+H]1+ | 648.6289 | -0.001 | 0.161085 | 0.172237 | 44.8 |
| POS | PC_37:5_[M+H]1+ / PE_40:5_[M+H]1+ / PA_42:6_[M+NH4]1+ | 794.5694 | -0.001 | 0.181068 | 0.192861 | 50.3 |
| NEG | PC-O_35:4_[M+Cl]1- | 788.5366 | 0.001 | 0.198949 | 0.211099 | 55.3 |
| NEG | PC-O_38:5_[M+Cl]1- | 828.5679 | 0.001 | 0.227041 | 0.23999 | 63.1 |
| POS | PC-P_40:6_[M+H]1+ | 818.6058 | -0.001 | 0.230458 | 0.242679 | 64.1 |
| NEG | PI_40:5_[M-H]1- | 911.5655 | 0.001 | 0.239531 | 0.251282 | 66.6 |
| POS | PS-O_38:4_[M+H]1+ / PG-O_38:6_[M+NH4]1+ | 798.5643 | 0.001 | 0.241801 | 0.25271 | 67.2 |
| NEG | FA(14:0)_[M-H]1- | 227.2017 | 0.001 | 0.266196 | 0.277163 | 74 |
| POS | CE_15:0_[M+NH4]1+ | 628.6027 | -0.001 | 0.31395 | 0.325664 | 87.3 |
| POS | PC_40:5_[M+H]1+ | 836.6164 | -0.001 | 0.355662 | 0.367562 | 98.9 |
| POS | CE_18:2_[M+NH4]1+ | 666.6184 | -0.001 | 0.400046 | 0.411899 | 111 |
| POS | CE_16:1_[M+NH4]1+ | 640.6027 | -0.001 | 0.44176 | 0.453171 | 123 |
| NEG | PC-O_38:4_[M+Cl]1- | 830.5836 | 0.000 | 0.518845 | 0.53029 | 144 |
| POS | SM_40:1_[M+H]1+ | 787.6688 | 0.000 | 0.623228 | 0.634642 | 173 |
| NEG | PE_37:1_[M-H]1- | 758.5705 | 0.000 | 0.694778 | 0.704921 | 193 |
| POS | PC_36:5_[M+H]1+ / PE_39:5_[M+H]1+ / PA_41:6_[M+NH4]1+ | 780.5538 | 0.000 | 0.776143 | 0.78461 | 216 |
| POS | PC_33:4_[M+H]1+ / PE_36:4_[M+H]1+ / PA_38:5_[M+NH4]1+ | 740.5225 | 0.000 | 0.82972 | 0.835732 | 231 |
| POS | CE_17:0_[M+NH4]1+ | 656.634 | 0.000 | 0.883292 | 0.88648 | 246 |
| POS | PC_35:2_[M+H]1+ / PE_38:2_[M+H]1+ / PA_40:3_[M+NH4]1+ | 772.5851 | 0.000 | 0.906085 | 0.906085 | 252 |

Supplementary Table 2. Lipid module assignment based on weighted correlation analysis

| Lipid | m/z | Module | Module color |
|---|---|---|---|
| LPE-P_16:0_[M-H]1- | 436.2833 | ME1 | black |
| PC-O-31:0_[M+OAC]1- | 764.5811 | ME1 | black |
| PC_37:3_[M+Cl]1- | 832.5629 | ME1 | black |
| PG-P_42:6_[M-H]1- | 833.5702 | ME1 | black |
| PC_39:4_[M+Cl]1- | 858.5785 | ME1 | black |
| PC-O_40:4_[M+Cl]1- | 858.6149 | ME1 | black |
| PC_39:3_[M+Cl]1- | 860.5942 | ME1 | black |
| PC_41:6_[M+Cl]1- | 882.5785 | ME1 | black |
| Hydroxycholesterol_[M+H-2H2O]1+ | 367.3359 | ME1 | black |
| PS-O_36:2_[M+H]1+ / PG-O_36:4_[M+NH4]1+ | 774.5643 | ME1 | black |
| PC-P_38:6_[M+H]1+ | 790.5745 | ME1 | black |
| PG_37:1_[M+H]1+ | 791.5797 | ME1 | black |
| PS-O_38:4_[M+H]1+ / PG-O_38:6_[M+NH4]1+ | 798.5643 | ME1 | black |
| PS-O_38:2_[M+H]1+ / PG-O_38:4_[M+NH4]1+ | 802.5956 | ME1 | black |
| PS-O_40:5_[M+H]1+ / PG-P_40:6_[M+NH4]1+ | 824.58 | ME1 | black |
| PC-2O_32:0_[M+H]1+ | 706.6109 | ME1 | black |
| FA(22:6)_[M-H]1- | 327.233 | ME2 | blue |
| PE-P_40:6_[M+H]1+ | 776.5589 | ME2 | blue |
| PG_36:1_[M+H]1+ | 777.564 | ME2 | blue |
| PC_36:5_[M+H]1+ / PE_39:5_[M+H]1+ / PA_41:6_[M+NH4]1+ | 780.5538 | ME2 | blue |
| PE-O_40:5_[M+H]1+ | 780.5902 | ME2 | blue |
| PC_36:4_[M+H]1+ / PE_39:4_[M+H]1+ / PA_41:5_[M+NH4]1+ | 782.5694 | ME2 | blue |
| PC_37:6_[M+H]1+ / PE_40:6_[M+H]1+ / PA_42:7_[M+NH4]1+ | 792.5538 | ME2 | blue |
| PC-O_38:6_[M+H]1+ | 792.5902 | ME2 | blue |
| PG_37:0_[M+H]1+ | 793.5953 | ME2 | blue |
| PC_37:5_[M+H]1+ / PE_40:5_[M+H]1+ / PA_42:6_[M+NH4]1+ | 794.5694 | ME2 | blue |
| PC-O_38:5_[M+H]1+ | 794.6058 | ME2 | blue |
| PC_37:4_[M+H]1+ / PE_40:4_[M+H]1+ / PA_42:5_[M+NH4]1+ | 796.5851 | ME2 | blue |
| PC-O_38:4_[M+H]1+ | 796.6215 | ME2 | blue |
| PC_38:6_[M+H]1+ / PE_41:6_[M+H]1+ | 806.5694 | ME2 | blue |
| PC_38:5_[M+H]1+ / PE_41:5_[M+H]1+ / PA_43:6_[M+NH4]1+ | 808.5851 | ME2 | blue |
| PC_38:4_[M+H]1+ / PE_41:4_[M+H]1+ | 810.6007 | ME2 | blue |
| PC_39:6_[M+H]1+ / PE_42:6_[M+H]1+ / PA_44:7_[M+NH4]1+ | 820.5851 | ME2 | blue |
| PC-O_40:6_[M+H]1+ | 820.6215 | ME2 | blue |
| PG_39:0_[M+H]1+ | 821.6266 | ME2 | blue |
| PC-O_40:4_[M+H]1+ | 824.6528 | ME2 | blue |
| PC_40:7_[M+H]1+ | 832.5851 | ME2 | blue |
| PC_40:6_[M+H]1+ / PE_43:6_[M+H]1+ | 834.6007 | ME2 | blue |
| PC_40:5_[M+H]1+ | 836.6164 | ME2 | blue |
| PC-P_42:4_[M+H]1+ | 850.6684 | ME2 | blue |
| PC-O_44:5_[M+H]1+ | 878.6997 | ME2 | blue |
| PI_38:4_[M+NH4]1+ | 904.591 | ME2 | blue |
| CE_20:5_[M+NH4]1+ | 688.6027 | ME2 | blue |
| CE_20:4_[M+NH4]1+ | 690.6184 | ME2 | blue |
| DG_40:3_[M+NH4]1+ | 692.6188 | ME2 | blue |
| CE_22:6_[M+NH4]1+ | 714.6184 | ME2 | blue |
| DG_42:5_[M+NH4]1+ | 716.6188 | ME2 | blue |

| | | | |
|---|---|---|---|
| PE-O_36:5_[M+H]1+ | 724.5276 | ME2 | blue |
| PE-P_38:6_[M+H]1+ | 748.5276 | ME2 | blue |
| PE-O_38:6_[M+H]1+ | 750.5432 | ME2 | blue |
| PG_34:0_[M+H]1+x | 751.5484 | ME2 | blue |
| PE-O_38:5_[M+H]1+ | 752.5589 | ME2 | blue |
| PC-O_36:5_[M+H]1+ | 766.5745 | ME2 | blue |
| PC-O_36:4_[M+H]1+ | 768.5902 | ME2 | blue |
| TG_49:2_[M+NH4]1+ | 834.7545 | ME3 | brown |
| TG_50:2_[M+NH4]1+ | 848.7702 | ME3 | brown |
| TG_50:1_[M+NH4]1+ | 850.7858 | ME3 | brown |
| TG_51:3_[M+NH4]1+ | 860.7702 | ME3 | brown |
| TG_51:2_[M+NH4]1+ | 862.7858 | ME3 | brown |
| TG_51:1_[M+NH4]1+ | 864.8015 | ME3 | brown |
| TG_52:4_[M+NH4]1+ | 872.7702 | ME3 | brown |
| TG_52:3_[M+NH4]1+ | 874.7858 | ME3 | brown |
| TG_52:2_[M+NH4]1+ | 876.8015 | ME3 | brown |
| TG_53:4_[M+NH4]1+ | 886.7858 | ME3 | brown |
| TG_53:3_[M+NH4]1+ | 888.8015 | ME3 | brown |
| TG_53:2_[M+NH4]1+ | 890.8171 | ME3 | brown |
| TG_54:5_[M+NH4]1+ | 898.7858 | ME3 | brown |
| TG_54:4_[M+NH4]1+ | 900.8015 | ME3 | brown |
| TG_54:3_[M+NH4]1+ | 902.8171 | ME3 | brown |
| TG_54:2_[M+NH4]1+ | 904.8328 | ME3 | brown |
| TG_54:1_[M+NH4]1+ | 906.8484 | ME3 | brown |
| TG_56:5_[M+NH4]1+ | 926.8171 | ME3 | brown |
| TG_56:4_[M+NH4]1+ | 928.8328 | ME3 | brown |
| DG_32:0_[M+H-H2O]1+ | 551.5034 | ME3 | brown |
| DG_34:2_[M+H-H2O]1+ | 575.5034 | ME3 | brown |
| DG_34:1_[M+H-H2O]1+ | 577.519 | ME3 | brown |
| DG_36:3_[M+H-H2O]1+ | 601.519 | ME3 | brown |
| DG_36:2_[M+H-H2O]1+ | 603.5347 | ME3 | brown |
| DG_36:1_[M+H-H2O]1+ | 605.5503 | ME3 | brown |
| DG_34:2_[M+NH4]1+ | 610.5405 | ME3 | brown |
| DG_34:1_[M+NH4]1+ | 612.5562 | ME3 | brown |
| DG_36:3_[M+NH4]1+ | 636.5562 | ME3 | brown |
| DG_36:2_[M+NH4]1+ | 638.5718 | ME3 | brown |
| PC_33:4_[M+H]1+ / PE_36:4_[M+H]1+ / PA_38:5_[M+NH4]1+ | 740.5225 | ME3 | brown |
| PC_35:4_[M+H]1+ / PE_38:4_[M+H]1+ / PA_40:5_[M+NH4]1+ | 768.5538 | ME3 | brown |
| SM_33:2_[M+OAC]1- | 745.5501 | ME4 | green |
| PG-O_35:1_[M-H]1- | 747.5545 | ME4 | green |
| PG-O_36:2_[M-H]1- | 759.5545 | ME4 | green |
| SM_34:1_[M+OAC]1- | 761.5814 | ME4 | green |
| PG-O_37:1_[M-H]1- | 775.5858 | ME4 | green |
| PG-O_38:2_[M-H]1- | 787.5858 | ME4 | green |
| PG-O_38:1_[M-H]1- | 789.6015 | ME4 | green |
| PC-O_35:2_[M+Cl]1- | 792.5679 | ME4 | green |
| PA_43:4_[M-H]1- | 793.5753 | ME4 | green |
| PC-O_36:4_[M+Cl]1- | 802.5523 | ME4 | green |
| PC-O_36:3_[M+Cl]1- | 804.5679 | ME4 | green |
| PA_44:5_[M-H]1- | 805.5753 | ME4 | green |

| | | | |
|---|---|---|---|
| PC-O_36:2_[M+Cl]1- | 806.5836 | ME4 | green |
| PG-O_40:2_[M-H]1- | 815.6171 | ME4 | green |
| PC_34:2_[M+OAC]1- / PS_38:1_[M-H]1- | 816.576 | ME4 | green |
| PG-O_40:1_[M-H]1- | 817.6328 | ME4 | green |
| PC_34:1_[M+OAC]1- / PS_38:0_[M-H]1- | 818.5917 | ME4 | green |
| PE_42:5_[M-H]1- | 820.5862 | ME4 | green |
| PC-O_38:5_[M+Cl]1- | 828.5679 | ME4 | green |
| PC-O_38:4_[M+Cl]1- | 830.5836 | ME4 | green |
| PG-P_41:0_[M-H]1- | 831.6484 | ME4 | green |
| PC_36:4_[M+OAC]1- / PS_40:3_[M-H]1- | 840.576 | ME4 | green |
| PG-O_42:2_[M-H]1- | 843.6484 | ME4 | green |
| PG-O_42:1_[M-H]1- | 845.6641 | ME4 | green |
| PE_44:5_[M-H]1- | 848.6175 | ME4 | green |
| PC-O_40:6_[M+Cl]1- | 854.5836 | ME4 | green |
| FA(14:1)_[M-H]1- | 225.186 | ME5 | grey |
| FA(20:1)_[M-H]1- | 309.2799 | ME5 | grey |
| OCT(C18H32O4)_[M-H]1- | 311.2228 | ME5 | grey |
| OCT(C18H34O4)_[M-H]1- | 313.2384 | ME5 | grey |
| FA(22:4)_[M-H]1- | 331.2643 | ME5 | grey |
| FA(22:1)_[M-H]1- | 337.3112 | ME5 | grey |
| EIC(C20H38O4)_[M-H]1- | 341.2697 | ME5 | grey |
| FA(14:0)_[M-H]1- | 227.2017 | ME5 | grey |
| BA(GDCA)_[M-H]1- | 448.3068 | ME5 | grey |
| FAHFA_PAHPA_[M-H]1- | 509.4575 | ME5 | grey |
| FA(16:1)_[M-H]1- | 253.2173 | ME5 | grey |
| LPC-O_16:1_[M+OAC]1- / LPS-O_20:0_[M-H]1- | 538.3514 | ME5 | grey |
| LPC-2O_16:0_[M+OAC]1- | 540.3671 | ME5 | grey |
| LPC_20:5_[M+OAC]1- | 600.3307 | ME5 | grey |
| FA(16:0)_[M-H]1- | 255.233 | ME5 | grey |
| PE_37:1_[M-H]1- | 758.5705 | ME5 | grey |
| FA(18:1)_[M-H]1- | 281.2486 | ME5 | grey |
| PA_43:6_[M-H]1- | 789.544 | ME5 | grey |
| FA(18:0)_[M-H]1- | 283.2643 | ME5 | grey |
| PC-P_36:5_[M+OAC]1- / PS-O_40:5_[M-H]1- | 822.5654 | ME5 | grey |
| FA(18:2)-OH_[M-H]1- | 295.2279 | ME5 | grey |
| FA(19:1)_[M-H]1- | 295.2643 | ME5 | grey |
| PI_40:5_[M-H]1- | 911.5655 | ME5 | grey |
| FA(18:1)-OH_[M-H]1- | 297.2435 | ME5 | grey |
| PI-P_30:1_[M+NH4]1+ | 782.5178 | ME5 | grey |
| PS_38:2_[M+H]1+ / PG_38:4_[M+NH4]1+ | 816.5749 | ME5 | grey |
| PG_44:6_[M+H]1+ | 879.611 | ME5 | grey |
| PI-O_38:3_[M+NH4]1+ | 892.6273 | ME5 | grey |
| PI_40:2_[M+NH4]1+ | 936.6536 | ME5 | grey |
| TG_58:10_[M+NH4]1+ | 944.7702 | ME5 | grey |
| MG_18:1_[M+NH4]1+ | 374.3265 | ME5 | grey |
| PE-O_34:3_[M+H]1+ | 700.5276 | ME5 | grey |
| PC_31:2_[M+H]1+ / PE_34:2_[M+H]1+ / PA_36:3_[M+NH4]1+ | 716.5225 | ME5 | grey |
| TG_52:6_[M+NH4]1+ | 868.7389 | ME6 | magenta |
| TG_52:5_[M+NH4]1+ | 870.7545 | ME6 | magenta |
| TG_54:7_[M+NH4]1+ | 894.7545 | ME6 | magenta |

| | | | |
|---|---|---|---|
| TG_54:6_[M+NH4]1+ | 896.7702 | ME6 | magenta |
| TG_56:9_[M+NH4]1+ | 918.7545 | ME6 | magenta |
| TG_56:8_[M+NH4]1+ | 920.7702 | ME6 | magenta |
| TG_56:7_[M+NH4]1+ | 922.7858 | ME6 | magenta |
| TG_56:6_[M+NH4]1+ | 924.8015 | ME6 | magenta |
| TG_58:9_[M+NH4]1+ | 946.7858 | ME6 | magenta |
| TG_58:8_[M+NH4]1+ | 948.8015 | ME6 | magenta |
| TG_58:7_[M+NH4]1+ | 950.8171 | ME6 | magenta |
| LPE_16:0_[M-H]1- | 452.2783 | ME7 | pink |
| LPE_18:0_[M-H]1- | 480.3096 | ME7 | pink |
| LPE_20:1_[M-H]1- | 506.3252 | ME7 | pink |
| LPE_20:0_[M-H]1- | 508.3409 | ME7 | pink |
| PC-O_16:0_[M+OAC]1- | 554.3463 | ME7 | pink |
| PC-O_16:0_[M+H]1+ / LPE_19:0_[M+H]1+ | 496.3398 | ME7 | pink |
| LPC-O_18:1_[M+H]1+ | 508.3762 | ME7 | pink |
| PC-O_18:1_[M+H]1+ | 522.3554 | ME7 | pink |
| PC-O_18:0_[M+H]1+ / LPE_21:0_[M+H]1+ | 524.3711 | ME7 | pink |
| LPC_20:4_[M+H]1+ | 544.3398 | ME7 | pink |
| LPC_20:3_[M+H]1+ | 546.3554 | ME7 | pink |
| LPC_13:0_[M+H]1+ / LPE_16:0_[M+H]1+ | 454.2928 | ME7 | pink |
| LPC-2O_16:0_[M+H]1+ | 482.3605 | ME7 | pink |
| LPC_16:1_[M+H]1+ / LPE_19:1_[M+H]1+ | 494.3241 | ME7 | pink |
| LPE_18:2_[M-H]1- | 476.2783 | ME8 | purple |
| LPE_18:1_[M-H]1- | 478.2939 | ME8 | purple |
| LPE_20:4_[M-H]1- | 500.2783 | ME8 | purple |
| LPE_20:2_[M-H]1- | 504.3096 | ME8 | purple |
| LPE_22:6_[M-H]1- | 524.2783 | ME8 | purple |
| LPC_18:2_[M+OAC]1- / LPS_22:1_[M-H]1- | 578.3463 | ME8 | purple |
| LPE_20:4_[M+H]1+ | 502.2928 | ME8 | purple |
| LPC_18:2_[M+H]1+ | 520.3398 | ME8 | purple |
| LPE_22:6_[M+H]1+ | 526.2928 | ME8 | purple |
| LPC_20:5_[M+H]1+ | 542.3241 | ME8 | purple |
| LPE_18:2_[M+H]1+ | 478.2928 | ME8 | purple |
| PC-P_35:2_[M+Cl]1- | 790.5523 | ME9 | red |
| PI_39:4_[M-H]1- | 899.5655 | ME9 | red |
| PC-O_36:2_[M+H]1+ / PE-P_39:1_[M+H]1+ | 772.6215 | ME9 | red |
| PC_35:1_[M+H]1+ / PE_38:1_[M+H]1+ / PA_40:2_[M+NH4]1+ | 774.6007 | ME9 | red |
| PC_36:6_[M+H]1+ / PE_39:6_[M+H]1+ / PA_41:7_[M+NH4]1+ | 778.5381 | ME9 | red |
| PC_36:3_[M+H]1+ / PE_39:3_[M+H]1+ / PA_41:4_[M+NH4]1+ | 784.5851 | ME9 | red |
| PC_36:2_[M+H]1+ / PE_39:2_[M+H]1+ / PA_41:3_[M+NH4]1+ | 786.6007 | ME9 | red |
| PC_36:1_[M+H]1+ / PE_39:1_[M+H]1+ / PA_41:2_[M+NH4]1+ | 788.6164 | ME9 | red |
| PC_38:3_[M+H]1+ / PE_41:3_[M+H]1+ / PA_43:4_[M+NH4]1+ | 812.6164 | ME9 | red |
| PC_38:2_[M+H]1+ / PE_41:2_[M+H]1+ | 814.632 | ME9 | red |
| PC_38:1_[M+H]1+ / PE_41:1_[M+H]1+ / PA_43:2_[M+NH4]1+ | 816.6477 | ME9 | red |
| PC_40:4_[M+H]1+ / PE_43:4_[M+H]1+ | 838.632 | ME9 | red |
| CE_18:3_[M+NH4]1+ | 664.6027 | ME9 | red |
| PC_32:1_[M+H]1+ / PE_35:1_[M+H]1+ / PA_37:2_[M+NH4]1+ | 732.5538 | ME9 | red |
| PC_33:3_[M+H]1+ / PE_36:3_[M+H]1+ / PA_38:4_[M+NH4]1+ | 742.5381 | ME9 | red |
| PC-O_34:3_[M+H]1+ / PE-P_37:2_[M+H]1+ | 742.5745 | ME9 | red |
| PC_33:2_[M+H]1+ / PE_36:2_[M+H]1+ / PA_38:3_[M+NH4]1+ | 744.5538 | ME9 | red |

| | | | |
|---|---|---|---|
| PC-O_34:2_[M+H]1+ / PE-O_37:2_[M+H]1+ | 744.5902 | ME9 | red |
| PC_33:1_[M+H]1+ / PE_36:1_[M+H]1+ / PA_38:2_[M+NH4]1+ | 746.5694 | ME9 | red |
| PC_34:4_[M+H]1+ / PE_37:4_[M+H]1+ / PA_39:5_[M+NH4]1+ | 754.5381 | ME9 | red |
| PC_34:3_[M+H]1+ / PE_37:3_[M+H]1+ / PA_39:4_[M+NH4]1+ | 756.5538 | ME9 | red |
| PC_34:2_[M+H]1+ / PE_37:2_[M+H]1+ / PA_39:3_[M+NH4]1+ | 758.5694 | ME9 | red |
| PC_34:1_[M+H]1+ / PE_37:1_[M+H]1+ / PA_39:2_[M+NH4]1+ | 760.5851 | ME9 | red |
| PC-O_36:3_[M+H]1+ | 770.6058 | ME9 | red |
| PC_35:2_[M+H]1+ / PE_38:2_[M+H]1+ / PA_40:3_[M+NH4]1+ | 772.5851 | ME9 | red |
| FA(20:4)_[M-H]1- | 303.233 | ME10 | turquoise |
| FA(24:1)_[M-H]1- | 365.3425 | ME10 | turquoise |
| SM_30:1_[M+OAC]1- | 705.5188 | ME10 | turquoise |
| SM_32:2_[M+OAC]1- | 731.5345 | ME10 | turquoise |
| PG-O_34:1_[M-H]1- | 733.5389 | ME10 | turquoise |
| SM_36:0_[M+OAC]1- | 791.6284 | ME10 | turquoise |
| SM_39:1_[M+H]1+ | 773.6531 | ME10 | turquoise |
| PE-O_40:6_[M+H]1+ | 778.5745 | ME10 | turquoise |
| SM_40:3_[M+H]1+ | 783.6375 | ME10 | turquoise |
| SM_40:2_[M+H]1+ | 785.6531 | ME10 | turquoise |
| SM_40:1_[M+H]1+ | 787.6688 | ME10 | turquoise |
| SM_41:1_[M+H]1+ | 799.6688 | ME10 | turquoise |
| SM_41:0_[M+H]1+ | 801.6844 | ME10 | turquoise |
| SM_42:3_[M+H]1+ | 811.6688 | ME10 | turquoise |
| SM_42:2_[M+H]1+ | 813.6844 | ME10 | turquoise |
| SM_42:1_[M+H]1+ | 815.7001 | ME10 | turquoise |
| PC-P_40:6_[M+H]1+ | 818.6058 | ME10 | turquoise |
| Cholesterol_[M+H-H2O]1+ | 369.3516 | ME10 | turquoise |
| CE_15:0_[M+NH4]1+ | 628.6027 | ME10 | turquoise |
| CE_16:1_[M+NH4]1+ | 640.6027 | ME10 | turquoise |
| CE_16:0_[M+NH4]1+ | 642.6184 | ME10 | turquoise |
| SM_30:1_[M+H]1+ | 647.5123 | ME10 | turquoise |
| Cer_42:2_[M+H]1+ | 648.6289 | ME10 | turquoise |
| Cer_42:1_[M+H]1+ | 650.6446 | ME10 | turquoise |
| CE_17:0_[M+NH4]1+ | 656.634 | ME10 | turquoise |
| CE_18:2_[M+NH4]1+ | 666.6184 | ME10 | turquoise |
| CE_18:1_[M+NH4]1+ | 668.634 | ME10 | turquoise |
| DG_38:0_[M+NH4]1+ | 670.6344 | ME10 | turquoise |
| SM_32:2_[M+H]1+ | 673.5279 | ME10 | turquoise |
| SM_32:1_[M+H]1+ | 675.5436 | ME10 | turquoise |
| DG_42:7_[M+H-H2O]1+ | 677.5503 | ME10 | turquoise |
| SM_33:1_[M+H]1+ | 689.5592 | ME10 | turquoise |
| SM_34:2_[M+H]1+ | 701.5592 | ME10 | turquoise |
| SM_34:1_[M+H]1+ | 703.5749 | ME10 | turquoise |
| DG_44:7_[M+H-H2O]1+ | 705.5816 | ME10 | turquoise |
| SM_35:1_[M+H]1+ | 717.5905 | ME10 | turquoise |
| PC-O_32:1_[M+H]1+ / PE-O_35:1_[M+H]1+ | 718.5745 | ME10 | turquoise |
| PC-O_32:0_[M+H]1+ / PE-O_35:0_[M+H]1+ | 720.5902 | ME10 | turquoise |
| SM_36:2_[M+H]1+ | 729.5905 | ME10 | turquoise |
| SM_36:1_[M+H]1+ | 731.6062 | ME10 | turquoise |
| PC_32:0_[M+H]1+ / PE_35:0_[M+H]1+ / PA_37:1_[M+NH4]1+ | 734.5694 | ME10 | turquoise |
| PC-O_34:1_[M+H]1+ / PE-O_37:1_[M+H]1+ | 746.6058 | ME10 | turquoise |

| | | | |
|---|---|---|---|
| SM_38:2_[M+H]1+ | 757.6218 | ME10 | turquoise |
| SM_38:1_[M+H]1+ | 759.6375 | ME10 | turquoise |
| PC-O_14:0_[M+OAC]1- | 526.315 | ME11 | yellow |
| PC-O_33:2_[M+Cl]1- | 764.5366 | ME11 | yellow |
| PA_41:4_[M-H]1- | 765.544 | ME11 | yellow |
| PC-O_35:4_[M+Cl]1- | 788.5366 | ME11 | yellow |
| DG_28:0_[M+H-H2O]1+ | 495.4408 | ME11 | yellow |
| TG_46:3_[M+NH4]1+ | 790.6919 | ME11 | yellow |
| TG_46:2_[M+NH4]1+ | 792.7076 | ME11 | yellow |
| TG_46:1_[M+NH4]1+ | 794.7232 | ME11 | yellow |
| TG_48:3_[M+NH4]1+ | 818.7232 | ME11 | yellow |
| TG_48:2_[M+NH4]1+ | 820.7389 | ME11 | yellow |
| DG_30:1_[M+H-H2O]1+ | 521.4564 | ME11 | yellow |
| TG_48:1_[M+NH4]1+ | 822.7545 | ME11 | yellow |
| TG_50:5_[M+NH4]1+ | 842.7232 | ME11 | yellow |
| TG_50:4_[M+NH4]1+ | 844.7389 | ME11 | yellow |
| TG_50:3_[M+NH4]1+ | 846.7545 | ME11 | yellow |
| DG_30:0_[M+H-H2O]1+ | 523.4721 | ME11 | yellow |
| TG_36:0_[M+NH4]1+ | 656.5824 | ME11 | yellow |
| TG_38:0_[M+NH4]1+ | 684.6137 | ME11 | yellow |
| DG_26:0_[M+H-H2O]1+ | 467.4095 | ME11 | yellow |
| TG_40:1_[M+NH4]1+ | 710.6293 | ME11 | yellow |
| TG_40:0_[M+NH4]1+ | 712.645 | ME11 | yellow |
| PC-O_14:0_[M+H]1+ / LPE_17:0_[M+H]1+ | 468.3085 | ME11 | yellow |
| DG_44:3_[M+H-H2O]1+ | 713.6442 | ME11 | yellow |
| TG_42:2_[M+NH4]1+ | 736.645 | ME11 | yellow |
| TG_42:1_[M+NH4]1+ | 738.6606 | ME11 | yellow |
| TG_42:0_[M+NH4]1+ | 740.6763 | ME11 | yellow |
| TG_44:2_[M+NH4]1+ | 764.6763 | ME11 | yellow |
| TG_44:1_[M+NH4]1+ | 766.6919 | ME11 | yellow |
| TG_44:0_[M+NH4]1+ | 768.7076 | ME11 | yellow |

Supplementary Table 3. Association between individual lipids and LAZ group over time

| Lipid | estimate for LAZ group | LAZ p value | LAZ FDR p value | LAZ*time p value | LAZ*time FDR p value |
|---|---|---|---|---|---|
| FA(14:1)_[M-H]1- | 0.039 | 0.498 | 0.932 | 0.452 | 0.999 |
| FA(14:0)_[M-H]1- | -0.016 | 0.785 | 0.932 | 0.985 | 0.999 |
| FA(16:1)_[M-H]1- | 0.014 | 0.803 | 0.932 | 0.706 | 0.999 |
| FA(16:0)_[M-H]1- | -0.016 | 0.782 | 0.932 | 0.637 | 0.999 |
| FA(18:1)_[M-H]1- | -0.010 | 0.865 | 0.954 | 0.441 | 0.999 |
| FA(18:0)_[M-H]1- | -0.044 | 0.447 | 0.932 | 0.882 | 0.999 |
| FA(18:2)-OH_[M-H]1- | 0.050 | 0.369 | 0.932 | 0.063 | 0.999 |
| FA(19:1)_[M-H]1- | -0.045 | 0.433 | 0.932 | 0.814 | 0.999 |
| FA(18:1)-OH_[M-H]1- | 0.048 | 0.359 | 0.932 | 0.071 | 0.999 |
| FA(20:4)_[M-H]1- | 0.057 | 0.324 | 0.932 | 0.063 | 0.999 |
| FA(20:1)_[M-H]1- | 0.013 | 0.824 | 0.943 | 0.080 | 0.999 |
| OCT(C18H32O4)_[M-H]1- | 0.015 | 0.779 | 0.932 | 0.104 | 0.999 |
| OCT(C18H34O4)_[M-H]1- | 0.052 | 0.334 | 0.932 | 0.013 | 0.999 |
| FA(22:6)_[M-H]1- | -0.016 | 0.786 | 0.932 | 0.842 | 0.999 |
| FA(22:4)_[M-H]1- | -0.001 | 0.985 | 0.994 | 0.649 | 0.999 |
| FA(22:1)_[M-H]1- | -0.133 | 0.021 | 0.932 | 0.175 | 0.999 |
| EIC(C20H38O4)_[M-H]1- | -0.015 | 0.792 | 0.932 | 0.732 | 0.999 |
| FA(24:1)_[M-H]1- | -0.113 | 0.045 | 0.932 | 0.320 | 0.999 |
| LPE-P_16:0_[M-H]1- | 0.011 | 0.849 | 0.952 | 0.945 | 0.999 |
| BA(GDCA)_[M-H]1- | -0.115 | 0.046 | 0.932 | 0.099 | 0.999 |
| LPE_16:0_[M-H]1- | -0.083 | 0.146 | 0.932 | 0.284 | 0.999 |
| LPE_18:2_[M-H]1- | -0.048 | 0.406 | 0.932 | 0.253 | 0.999 |
| LPE_18:1_[M-H]1- | -0.147 | 0.011 | 0.932 | 0.050 | 0.999 |
| LPE_18:0_[M-H]1- | -0.108 | 0.051 | 0.932 | 0.353 | 0.999 |
| LPE_20:4_[M-H]1- | -0.054 | 0.331 | 0.932 | 0.395 | 0.999 |
| LPE_20:2_[M-H]1- | -0.085 | 0.126 | 0.932 | 0.188 | 0.999 |
| LPE_20:1_[M-H]1- | -0.125 | 0.024 | 0.932 | 0.168 | 0.999 |
| LPE_20:0_[M-H]1- | -0.093 | 0.079 | 0.932 | 0.430 | 0.999 |
| FAHFA_PAHPA_[M-H]1- | 0.005 | 0.927 | 0.965 | 0.793 | 0.999 |
| LPE_22:6_[M-H]1- | -0.107 | 0.060 | 0.932 | 0.038 | 0.999 |
| PC-O_14:0_[M+OAC]1- | -0.020 | 0.720 | 0.932 | 0.989 | 0.999 |
| LPC-O_16:1_[M+OAC]1- / LPS-O_20:0_[M-H]1- | -0.006 | 0.914 | 0.965 | 0.853 | 0.999 |
| LPC-2O_16:0_[M+OAC]1- | 0.061 | 0.282 | 0.932 | 0.431 | 0.999 |
| PC-O_16:0_[M+OAC]1- | -0.026 | 0.631 | 0.932 | 0.647 | 0.999 |
| LPC_18:2_[M+OAC]1- / LPS_22:1_[M-H]1- | 0.018 | 0.743 | 0.932 | 0.915 | 0.999 |
| LPC_20:5_[M+OAC]1- | -0.007 | 0.902 | 0.965 | 0.808 | 0.999 |
| SM_30:1_[M+OAC]1- | -0.011 | 0.833 | 0.949 | 0.856 | 0.999 |
| SM_32:2_[M+OAC]1- | -0.010 | 0.844 | 0.952 | 0.787 | 0.999 |
| PG-O_34:1_[M-H]1- | -0.015 | 0.748 | 0.932 | 0.960 | 0.999 |
| SM_33:2_[M+OAC]1- | -0.133 | 0.019 | 0.932 | 0.424 | 0.999 |
| PG-O_35:1_[M-H]1- | -0.032 | 0.585 | 0.932 | 0.899 | 0.999 |
| PE_37:1_[M-H]1- | -0.041 | 0.487 | 0.932 | 0.154 | 0.999 |
| PG-O_36:2_[M-H]1- | -0.036 | 0.523 | 0.932 | 0.683 | 0.999 |
| SM_34:1_[M+OAC]1- | -0.089 | 0.123 | 0.932 | 0.184 | 0.999 |
| PC-O_33:2_[M+Cl]1- | -0.060 | 0.282 | 0.932 | 0.481 | 0.999 |
| PC-O-31:0_[M+OAC]1- | -0.006 | 0.917 | 0.965 | 0.943 | 0.999 |
| PA_41:4_[M-H]1- | -0.065 | 0.242 | 0.932 | 0.418 | 0.999 |
| PG-O_37:1_[M-H]1- | -0.065 | 0.236 | 0.932 | 0.343 | 0.999 |
| PG-O_38:2_[M-H]1- | -0.028 | 0.600 | 0.932 | 0.985 | 0.999 |
| PC-O_35:4_[M+Cl]1- | -0.042 | 0.471 | 0.932 | 0.851 | 0.999 |
| PA_43:6_[M-H]1- | -0.016 | 0.788 | 0.932 | 0.545 | 0.999 |
| PG-O_38:1_[M-H]1- | -0.049 | 0.322 | 0.932 | 0.884 | 0.999 |
| PC-P_35:2_[M+Cl]1- | -0.056 | 0.325 | 0.932 | 0.694 | 0.999 |
| SM_36:0_[M+OAC]1- | 0.023 | 0.675 | 0.932 | 0.093 | 0.999 |
| PC-O_35:2_[M+Cl]1- | -0.071 | 0.213 | 0.932 | 0.867 | 0.999 |
| PA_43:4_[M-H]1- | -0.084 | 0.143 | 0.932 | 0.685 | 0.999 |
| PC-O_36:4_[M+Cl]1- | -0.034 | 0.568 | 0.932 | 0.998 | 0.999 |
| PC-O_36:3_[M+Cl]1- | -0.022 | 0.700 | 0.932 | 0.947 | 0.999 |
| PA_44:5_[M-H]1- | -0.015 | 0.803 | 0.932 | 0.980 | 0.999 |
| PC-O_36:2_[M+Cl]1- | -0.054 | 0.353 | 0.932 | 0.929 | 0.999 |
| PG-O_40:2_[M-H]1- | -0.038 | 0.490 | 0.932 | 0.817 | 0.999 |
| PC_34:2_[M+OAC]1- / PS_38:1_[M-H]1- | -0.093 | 0.110 | 0.932 | 0.044 | 0.999 |
| PG-O_40:1_[M-H]1- | -0.068 | 0.171 | 0.932 | 0.704 | 0.999 |
| PC_34:1_[M+OAC]1- / PS_38:0_[M-H]1- | -0.108 | 0.058 | 0.932 | 0.079 | 0.999 |
| PE_42:5_[M-H]1- | -0.058 | 0.274 | 0.932 | 0.992 | 0.999 |
| PC-P_36:5_[M+OAC]1- / PS-O_40:5_[M-H]1- | 0.028 | 0.636 | 0.932 | 0.579 | 0.999 |
| PC-O_38:5_[M+Cl]1- | -0.059 | 0.306 | 0.932 | 0.327 | 0.999 |
| PC-O_38:4_[M+Cl]1- | -0.101 | 0.077 | 0.932 | 0.256 | 0.999 |
| PG-P_41:0_[M-H]1- | -0.035 | 0.525 | 0.932 | 0.751 | 0.999 |
| PC_37:3_[M+Cl]1- | 0.007 | 0.895 | 0.965 | 0.892 | 0.999 |
| PG-P_42:6_[M-H]1- | -0.001 | 0.993 | 0.996 | 0.591 | 0.999 |
| PC_36:4_[M+OAC]1- / PS_40:3_[M-H]1- | 0.005 | 0.932 | 0.965 | 0.422 | 0.999 |
| PG-O_42:2_[M-H]1- | -0.043 | 0.426 | 0.932 | 0.758 | 0.999 |
| PG-O_42:1_[M-H]1- | -0.069 | 0.233 | 0.932 | 0.955 | 0.999 |
| PE_44:5_[M-H]1- | -0.161 | 0.006 | 0.932 | 0.060 | 0.999 |
| PC-O_40:6_[M+Cl]1- | -0.076 | 0.185 | 0.932 | 0.604 | 0.999 |
| PC_39:4_[M+Cl]1- | -0.017 | 0.774 | 0.932 | 0.519 | 0.999 |
| PC-O_40:4_[M+Cl]1- | -0.019 | 0.747 | 0.932 | 0.772 | 0.999 |
| PC_39:3_[M+Cl]1- | -0.021 | 0.710 | 0.932 | 0.270 | 0.999 |
| PC_41:6_[M+Cl]1- | -0.041 | 0.474 | 0.932 | 0.315 | 0.999 |
| PI_39:4_[M-H]1- | 0.041 | 0.479 | 0.932 | 0.479 | 0.999 |
| PI_40:5_[M-H]1- | -0.102 | 0.078 | 0.932 | 0.092 | 0.999 |
| Hydroxycholesterol_[M+H-2H2O]1+ | -0.017 | 0.772 | 0.932 | 0.428 | 0.999 |
| Cholesterol_[M+H-H2O]1+ | -0.091 | 0.115 | 0.932 | 0.395 | 0.999 |
| MG_18:1_[M+NH4]1+ | 0.100 | 0.072 | 0.932 | 0.159 | 0.999 |
| LPC_13:0_[M+H]1+ / LPE_16:0_[M+H]1+ | -0.010 | 0.866 | 0.954 | 0.719 | 0.999 |
| DG_26:0_[M+H-H2O]1+ | 0.020 | 0.724 | 0.932 | 0.478 | 0.999 |
| PC-O_14:0_[M+H]1+ / LPE_17:0_[M+H]1+ | -0.027 | 0.624 | 0.932 | 0.935 | 0.999 |
| LPE_18:2_[M+H]1+ | -0.022 | 0.699 | 0.932 | 0.329 | 0.999 |
| LPC-2O_16:0_[M+H]1+ | 0.022 | 0.697 | 0.932 | 0.730 | 0.999 |

| | | | | | |
|---|---|---|---|---|---|
| LPC_16:1_[M+H]1+ / LPE_19:1_[M+H]1+ | -0.036 | 0.533 | 0.932 | 0.903 | 0.999 |
| DG_28:0_[M+H-H2O]1+ | -0.019 | 0.729 | 0.932 | 0.694 | 0.999 |
| PC-O_16:0_[M+H]1+ / LPE_19:0_[M+H]1+ | 0.001 | 0.987 | 0.994 | 0.734 | 0.999 |
| LPE_20:4_[M+H]1+ | -0.029 | 0.603 | 0.932 | 0.406 | 0.999 |
| LPC-O_18:1_[M+H]1+ | -0.009 | 0.870 | 0.954 | 0.822 | 0.999 |
| LPC_18:2_[M+H]1+ | -0.020 | 0.720 | 0.932 | 0.590 | 0.999 |
| DG_30:1_[M+H-H2O]1+ | -0.005 | 0.926 | 0.965 | 0.876 | 0.999 |
| PC-O_18:1_[M+H]1+ | -0.049 | 0.368 | 0.932 | 0.517 | 0.999 |
| DG_30:0_[M+H-H2O]1+ | -0.023 | 0.678 | 0.932 | 0.670 | 0.999 |
| PC-O_18:0_[M+H]1+ / LPE_21:0_[M+H]1+ | -0.024 | 0.644 | 0.932 | 0.896 | 0.999 |
| LPE_22:6_[M+H]1+ | -0.039 | 0.486 | 0.932 | 0.309 | 0.999 |
| LPC_20:5_[M+H]1+ | 0.040 | 0.482 | 0.932 | 0.669 | 0.999 |
| LPC_20:4_[M+H]1+ | -0.010 | 0.844 | 0.952 | 0.926 | 0.999 |
| LPC_20:3_[M+H]1+ | 0.034 | 0.519 | 0.932 | 0.232 | 0.999 |
| DG_32:0_[M+H-H2O]1+ | -0.035 | 0.543 | 0.932 | 0.590 | 0.999 |
| DG_34:2_[M+H-H2O]1+ | -0.028 | 0.624 | 0.932 | 0.954 | 0.999 |
| DG_34:1_[M+H-H2O]1+ | -0.021 | 0.722 | 0.932 | 0.891 | 0.999 |
| DG_36:3_[M+H-H2O]1+ | -0.032 | 0.576 | 0.932 | 0.798 | 0.999 |
| DG_36:2_[M+H-H2O]1+ | -0.023 | 0.689 | 0.932 | 0.764 | 0.999 |
| DG_36:1_[M+H-H2O]1+ | -0.034 | 0.554 | 0.932 | 0.926 | 0.999 |
| DG_34:2_[M+NH4]1+ | 0.095 | 0.093 | 0.932 | 0.009 | 0.999 |
| DG_34:1_[M+NH4]1+ | 0.089 | 0.107 | 0.932 | 0.110 | 0.999 |
| CE_15:0_[M+NH4]1+ | -0.008 | 0.891 | 0.965 | 0.447 | 0.999 |
| DG_36:3_[M+NH4]1+ | 0.060 | 0.301 | 0.932 | 0.122 | 0.999 |
| DG_36:2_[M+NH4]1+ | 0.046 | 0.428 | 0.932 | 0.092 | 0.999 |
| CE_16:1_[M+NH4]1+ | 0.035 | 0.537 | 0.932 | 0.916 | 0.999 |
| CE_16:0_[M+NH4]1+ | -0.014 | 0.801 | 0.932 | 0.959 | 0.999 |
| SM_30:1_[M+H]1+ | -0.050 | 0.319 | 0.932 | 0.356 | 0.999 |
| Cer_42:2_[M+H]1+ | 0.017 | 0.774 | 0.932 | 0.854 | 0.999 |
| Cer_42:1_[M+H]1+ | 0.011 | 0.849 | 0.952 | 0.934 | 0.999 |
| TG_36:0_[M+NH4]1+ | 0.073 | 0.196 | 0.932 | 0.395 | 0.999 |
| CE_17:0_[M+NH4]1+ | 0.032 | 0.581 | 0.932 | 0.889 | 0.999 |
| CE_18:3_[M+NH4]1+ | -0.003 | 0.958 | 0.979 | 0.869 | 0.999 |
| CE_18:2_[M+NH4]1+ | -0.042 | 0.473 | 0.932 | 0.543 | 0.999 |
| CE_18:1_[M+NH4]1+ | -0.026 | 0.654 | 0.932 | 0.607 | 0.999 |
| DG_38:0_[M+NH4]1+ | -0.017 | 0.771 | 0.932 | 0.937 | 0.999 |
| SM_32:2_[M+H]1+ | -0.039 | 0.494 | 0.932 | 0.285 | 0.999 |
| SM_32:1_[M+H]1+ | -0.034 | 0.470 | 0.932 | 0.817 | 0.999 |
| DG_42:7_[M+H-H2O]1+ | -0.082 | 0.115 | 0.932 | 0.098 | 0.999 |
| TG_38:0_[M+NH4]1+ | 0.047 | 0.398 | 0.932 | 0.775 | 0.999 |
| CE_20:5_[M+NH4]1+ | -0.028 | 0.631 | 0.932 | 0.773 | 0.999 |
| SM_33:1_[M+H]1+ | -0.026 | 0.648 | 0.932 | 0.523 | 0.999 |
| CE_20:4_[M+NH4]1+ | -0.033 | 0.549 | 0.932 | 0.870 | 0.999 |
| DG_40:3_[M+NH4]1+ | -0.045 | 0.412 | 0.932 | 0.911 | 0.999 |
| PE-O_34:3_[M+H]1+ | 0.028 | 0.619 | 0.932 | 0.858 | 0.999 |
| SM_34:2_[M+H]1+ | -0.041 | 0.467 | 0.932 | 0.911 | 0.999 |
| SM_34:1_[M+H]1+ | -0.063 | 0.275 | 0.932 | 0.817 | 0.999 |
| DG_44:7_[M+H-H2O]1+ | -0.048 | 0.403 | 0.932 | 0.998 | 0.999 |
| PC-2O_32:0_[M+H]1+ | 0.002 | 0.977 | 0.991 | 0.990 | 0.999 |
| TG_40:1_[M+NH4]1+ | 0.068 | 0.215 | 0.932 | 0.357 | 0.999 |
| TG_40:0_[M+NH4]1+ | 0.016 | 0.773 | 0.932 | 0.986 | 0.999 |
| DG_44:3_[M+H-H2O]1+ | 0.019 | 0.730 | 0.932 | 0.953 | 0.999 |
| CE_22:6_[M+NH4]1+ | -0.049 | 0.392 | 0.932 | 0.832 | 0.999 |
| PC_31:2_[M+H]1+ / PE_34:2_[M+H]1+ / PA_36:3_[M+NH4]1+ | 0.007 | 0.905 | 0.965 | 0.986 | 0.999 |
| DG_42:5_[M+NH4]1+ | -0.060 | 0.295 | 0.932 | 0.344 | 0.999 |
| SM_35:1_[M+H]1+ | -0.048 | 0.389 | 0.932 | 0.565 | 0.999 |
| PC-O_32:1_[M+H]1+ / PE-O_35:1_[M+H]1+ | -0.016 | 0.788 | 0.932 | 0.866 | 0.999 |
| PC-O_32:0_[M+H]1+ / PE-O_35:0_[M+H]1+ | -0.048 | 0.413 | 0.932 | 0.534 | 0.999 |
| PE-O_36:5_[M+H]1+ | -0.010 | 0.862 | 0.954 | 0.790 | 0.999 |
| SM_36:2_[M+H]1+ | -0.040 | 0.454 | 0.932 | 0.645 | 0.999 |
| SM_36:1_[M+H]1+ | -0.039 | 0.432 | 0.932 | 0.986 | 0.999 |
| PC_32:1_[M+H]1+ / PE_35:1_[M+H]1+ / PA_37:2_[M+NH4]1+ | -0.037 | 0.520 | 0.932 | 0.658 | 0.999 |
| PC_32:0_[M+H]1+ / PE_35:0_[M+H]1+ / PA_37:1_[M+NH4]1+ | -0.043 | 0.448 | 0.932 | 0.977 | 0.999 |
| TG_42:2_[M+NH4]1+ | 0.077 | 0.157 | 0.932 | 0.370 | 0.999 |
| TG_42:1_[M+NH4]1+ | 0.075 | 0.167 | 0.932 | 0.224 | 0.999 |
| PC_33:4_[M+H]1+ / PE_36:4_[M+H]1+ / PA_38:5_[M+NH4]1+ | 0.046 | 0.442 | 0.932 | 0.066 | 0.999 |
| TG_42:0_[M+NH4]1+ | -0.006 | 0.918 | 0.965 | 0.861 | 0.999 |
| PC_33:3_[M+H]1+ / PE_36:3_[M+H]1+ / PA_38:4_[M+NH4]1+ | -0.036 | 0.531 | 0.932 | 0.740 | 0.999 |
| PC-O_34:3_[M+H]1+ / PE-P_37:2_[M+H]1+ | -0.004 | 0.943 | 0.968 | 0.558 | 0.999 |
| PC_33:2_[M+H]1+ / PE_36:2_[M+H]1+ / PA_38:3_[M+NH4]1+ | -0.017 | 0.770 | 0.932 | 0.841 | 0.999 |
| PC-O_34:2_[M+H]1+ / PE-O_37:2_[M+H]1+ | 0.022 | 0.695 | 0.932 | 0.294 | 0.999 |
| PC_33:1_[M+H]1+ / PE_36:1_[M+H]1+ / PA_38:2_[M+NH4]1+ | -0.069 | 0.221 | 0.932 | 0.160 | 0.999 |
| PC-O_34:1_[M+H]1+ / PE-O_37:1_[M+H]1+ | -0.026 | 0.661 | 0.932 | 0.847 | 0.999 |
| PE-P_38:6_[M+H]1+ | 0.002 | 0.973 | 0.991 | 0.633 | 0.999 |
| PE-O_38:6_[M+H]1+ | -0.023 | 0.648 | 0.932 | 0.912 | 0.999 |
| PG_34:0_[M+H]1+x | -0.022 | 0.651 | 0.932 | 0.913 | 0.999 |
| PE-O_38:5_[M+H]1+ | -0.036 | 0.513 | 0.932 | 0.663 | 0.999 |
| PC_34:4_[M+H]1+ / PE_37:4_[M+H]1+ / PA_39:5_[M+NH4]1+ | -0.042 | 0.469 | 0.932 | 0.738 | 0.999 |
| PC_34:3_[M+H]1+ / PE_37:3_[M+H]1+ / PA_39:4_[M+NH4]1+ | -0.005 | 0.936 | 0.965 | 0.754 | 0.999 |
| SM_38:2_[M+H]1+ | -0.026 | 0.644 | 0.932 | 0.907 | 0.999 |
| PC_34:2_[M+H]1+ / PE_37:2_[M+H]1+ / PA_39:3_[M+NH4]1+ | -0.047 | 0.419 | 0.932 | 0.503 | 0.999 |
| SM_38:1_[M+H]1+ | -0.045 | 0.368 | 0.932 | 0.852 | 0.999 |
| PC_34:1_[M+H]1+ / PE_37:1_[M+H]1+ / PA_39:2_[M+NH4]1+ | -0.064 | 0.258 | 0.932 | 0.430 | 0.999 |
| TG_44:2_[M+NH4]1+ | 0.043 | 0.425 | 0.932 | 0.676 | 0.999 |
| PC-O_36:5_[M+H]1+ | -0.014 | 0.805 | 0.932 | 0.552 | 0.999 |
| TG_44:1_[M+NH4]1+ | 0.042 | 0.437 | 0.932 | 0.627 | 0.999 |
| PC_35:4_[M+H]1+ / PE_38:4_[M+H]1+ / PA_40:5_[M+NH4]1+ | -0.015 | 0.797 | 0.932 | 0.311 | 0.999 |
| PC-O_36:4_[M+H]1+ | -0.010 | 0.865 | 0.954 | 0.592 | 0.999 |
| TG_44:0_[M+NH4]1+ | -0.014 | 0.803 | 0.932 | 0.845 | 0.999 |
| PC-O_36:3_[M+H]1+ | -0.044 | 0.453 | 0.932 | 0.539 | 0.999 |
| PC_35:2_[M+H]1+ / PE_38:2_[M+H]1+ / PA_40:3_[M+NH4]1+ | -0.052 | 0.373 | 0.932 | 0.386 | 0.999 |
| PC-O_36:2_[M+H]1+ / PE-P_39:1_[M+H]1+ | -0.029 | 0.619 | 0.932 | 0.485 | 0.999 |
| SM_39:1_[M+H]1+ | -0.028 | 0.592 | 0.932 | 0.790 | 0.999 |

| | | | | | |
|---|---|---|---|---|---|
| PS-O_36:2_[M+H]1+ / PG-O_36:4_[M+NH4]1+ | -0.009 | 0.875 | 0.954 | 0.429 | 0.999 |
| PC_35:1_[M+H]1+ / PE_38:1_[M+H]1+ / PA_40:2_[M+NH4]1+ | -0.075 | 0.188 | 0.932 | 0.234 | 0.999 |
| PE-P_40:6_[M+H]1+ | -0.022 | 0.701 | 0.932 | 0.917 | 0.999 |
| PG_36:1_[M+H]1+ | 0.065 | 0.248 | 0.932 | 0.040 | 0.999 |
| PC_36:6_[M+H]1+ / PE_39:6_[M+H]1+ / PA_41:7_[M+NH4]1+ | 0.023 | 0.689 | 0.932 | 0.207 | 0.999 |
| PE-O_40:6_[M+H]1+ | 0.026 | 0.648 | 0.932 | 0.215 | 0.999 |
| PC_36:5_[M+H]1+ / PE_39:5_[M+H]1+ / PA_41:6_[M+NH4]1+ | 0.000 | 0.996 | 0.996 | 0.729 | 0.999 |
| PE-O_40:5_[M+H]1+ | -0.072 | 0.208 | 0.932 | 0.061 | 0.999 |
| PI-P_30:1_[M+NH4]1+ | -0.020 | 0.724 | 0.932 | 0.991 | 0.999 |
| PC_36:4_[M+H]1+ / PE_39:4_[M+H]1+ / PA_41:5_[M+NH4]1+ | -0.023 | 0.683 | 0.932 | 0.664 | 0.999 |
| SM_40:3_[M+H]1+ | -0.064 | 0.202 | 0.932 | 0.630 | 0.999 |
| PC_36:3_[M+H]1+ / PE_39:3_[M+H]1+ / PA_41:4_[M+NH4]1+ | -0.076 | 0.193 | 0.932 | 0.563 | 0.999 |
| SM_40:2_[M+H]1+ | -0.045 | 0.414 | 0.932 | 0.932 | 0.999 |
| PC_36:2_[M+H]1+ / PE_39:2_[M+H]1+ / PA_41:3_[M+NH4]1+ | -0.082 | 0.157 | 0.932 | 0.262 | 0.999 |
| SM_40:1_[M+H]1+ | -0.058 | 0.327 | 0.932 | 0.785 | 0.999 |
| PC_36:1_[M+H]1+ / PE_39:1_[M+H]1+ / PA_41:2_[M+NH4]1+ | -0.108 | 0.065 | 0.932 | 0.215 | 0.999 |
| PC-P_38:6_[M+H]1+ | -0.020 | 0.725 | 0.932 | 0.332 | 0.999 |
| TG_46:3_[M+NH4]1+ | 0.057 | 0.290 | 0.932 | 0.347 | 0.999 |
| PG_37:1_[M+H]1+ | -0.016 | 0.782 | 0.932 | 0.531 | 0.999 |
| PC_37:6_[M+H]1+ / PE_40:6_[M+H]1+ / PA_42:7_[M+NH4]1+ | 0.009 | 0.871 | 0.954 | 0.673 | 0.999 |
| PC-O_38:6_[M+H]1+ | -0.005 | 0.938 | 0.965 | 0.534 | 0.999 |
| TG_46:2_[M+NH4]1+ | 0.032 | 0.558 | 0.932 | 0.713 | 0.999 |
| PG_37:0_[M+H]1+ | -0.007 | 0.899 | 0.965 | 0.637 | 0.999 |
| PC_37:5_[M+H]1+ / PE_40:5_[M+H]1+ / PA_42:6_[M+NH4]1+ | 0.031 | 0.586 | 0.932 | 0.322 | 0.999 |
| PC-O_38:5_[M+H]1+ | -0.026 | 0.646 | 0.932 | 0.982 | 0.999 |
| TG_46:1_[M+NH4]1+ | 0.027 | 0.615 | 0.932 | 0.752 | 0.999 |
| PC_37:4_[M+H]1+ / PE_40:4_[M+H]1+ / PA_42:5_[M+NH4]1+ | -0.042 | 0.467 | 0.932 | 0.861 | 0.999 |
| PC-O_38:4_[M+H]1+ | -0.006 | 0.921 | 0.965 | 0.803 | 0.999 |
| PS-O_38:4_[M+H]1+ / PG-O_38:6_[M+NH4]1+ | -0.041 | 0.481 | 0.932 | 0.444 | 0.999 |
| SM_41:1_[M+H]1+ | -0.039 | 0.493 | 0.932 | 0.735 | 0.999 |
| SM_41:0_[M+H]1+ | -0.041 | 0.485 | 0.932 | 0.769 | 0.999 |
| PS-O_38:2_[M+H]1+ / PG-O_38:4_[M+NH4]1+ | -0.027 | 0.647 | 0.932 | 0.305 | 0.999 |
| PC_38:6_[M+H]1+ / PE_41:6_[M+H]1+ | -0.030 | 0.603 | 0.932 | 0.884 | 0.999 |
| PC_38:5_[M+H]1+ / PE_41:5_[M+H]1+ / PA_43:6_[M+NH4]1+ | -0.037 | 0.524 | 0.932 | 0.715 | 0.999 |
| PC_38:4_[M+H]1+ / PE_41:4_[M+H]1+ | -0.040 | 0.452 | 0.932 | 0.890 | 0.999 |
| SM_42:3_[M+H]1+ | -0.055 | 0.300 | 0.932 | 0.993 | 0.999 |
| PC_38:3_[M+H]1+ / PE_41:3_[M+H]1+ / PA_43:4_[M+NH4]1+ | -0.091 | 0.092 | 0.932 | 0.653 | 0.999 |
| SM_42:2_[M+H]1+ | -0.060 | 0.283 | 0.932 | 0.999 | 0.999 |
| PC_38:2_[M+H]1+ / PE_41:2_[M+H]1+ | -0.106 | 0.063 | 0.932 | 0.126 | 0.999 |
| SM_42:1_[M+H]1+ | -0.048 | 0.414 | 0.932 | 0.916 | 0.999 |
| PS_38:2_[M+H]1+ / PG_38:4_[M+NH4]1+ | -0.052 | 0.368 | 0.932 | 0.583 | 0.999 |
| PC_38:1_[M+H]1+ / PE_41:1_[M+H]1+ / PA_43:2_[M+NH4]1+ | -0.028 | 0.631 | 0.932 | 0.982 | 0.999 |
| PC-P_40:6_[M+H]1+ | -0.048 | 0.406 | 0.932 | 0.307 | 0.999 |
| TG_48:3_[M+NH4]1+ | 0.052 | 0.330 | 0.932 | 0.461 | 0.999 |
| PC_39:6_[M+H]1+ / PE_42:6_[M+H]1+ / PA_44:7_[M+NH4]1+ | -0.016 | 0.781 | 0.932 | 0.877 | 0.999 |
| PC-O_40:6_[M+H]1+ | -0.014 | 0.817 | 0.943 | 0.944 | 0.999 |
| TG_48:2_[M+NH4]1+ | 0.042 | 0.444 | 0.932 | 0.600 | 0.999 |
| PG_39:0_[M+H]1+ | -0.025 | 0.665 | 0.932 | 0.799 | 0.999 |
| TG_48:1_[M+NH4]1+ | 0.023 | 0.684 | 0.932 | 0.792 | 0.999 |
| PS-O_40:5_[M+H]1+ / PG-P_40:6_[M+NH4]1+ | -0.072 | 0.215 | 0.932 | 0.153 | 0.999 |
| PC-O_40:4_[M+H]1+ | -0.035 | 0.557 | 0.932 | 0.694 | 0.999 |
| PC_40:7_[M+H]1+ | 0.006 | 0.913 | 0.965 | 0.196 | 0.999 |
| PC_40:6_[M+H]1+ / PE_43:6_[M+H]1+ | -0.068 | 0.217 | 0.932 | 0.752 | 0.999 |
| TG_49:2_[M+NH4]1+ | 0.037 | 0.530 | 0.932 | 0.590 | 0.999 |
| PC_40:5_[M+H]1+ | -0.107 | 0.066 | 0.932 | 0.112 | 0.999 |
| PC_40:4_[M+H]1+ / PE_43:4_[M+H]1+ | -0.070 | 0.232 | 0.932 | 0.553 | 0.999 |
| TG_50:5_[M+NH4]1+ | 0.037 | 0.502 | 0.932 | 0.521 | 0.999 |
| TG_50:4_[M+NH4]1+ | 0.053 | 0.337 | 0.932 | 0.472 | 0.999 |
| TG_50:3_[M+NH4]1+ | 0.057 | 0.309 | 0.932 | 0.434 | 0.999 |
| TG_50:2_[M+NH4]1+ | 0.051 | 0.376 | 0.932 | 0.546 | 0.999 |
| PC-P_42:4_[M+H]1+ | -0.046 | 0.437 | 0.932 | 0.508 | 0.999 |
| TG_50:1_[M+NH4]1+ | 0.022 | 0.708 | 0.932 | 0.991 | 0.999 |
| TG_51:3_[M+NH4]1+ | 0.016 | 0.788 | 0.932 | 0.892 | 0.999 |
| TG_51:2_[M+NH4]1+ | 0.036 | 0.539 | 0.932 | 0.530 | 0.999 |
| TG_51:1_[M+NH4]1+ | 0.031 | 0.595 | 0.932 | 0.591 | 0.999 |
| TG_52:6_[M+NH4]1+ | 0.033 | 0.564 | 0.932 | 0.537 | 0.999 |
| TG_52:5_[M+NH4]1+ | 0.037 | 0.519 | 0.932 | 0.725 | 0.999 |
| TG_52:4_[M+NH4]1+ | 0.039 | 0.502 | 0.932 | 0.896 | 0.999 |
| TG_52:3_[M+NH4]1+ | 0.052 | 0.376 | 0.932 | 0.644 | 0.999 |
| TG_52:2_[M+NH4]1+ | 0.052 | 0.382 | 0.932 | 0.415 | 0.999 |
| PC-O_44:5_[M+H]1+ | -0.051 | 0.371 | 0.932 | 0.826 | 0.999 |
| PG_44:6_[M+H]1+ | 0.019 | 0.731 | 0.932 | 0.260 | 0.999 |
| TG_53:4_[M+NH4]1+ | 0.025 | 0.664 | 0.932 | 0.937 | 0.999 |
| TG_53:3_[M+NH4]1+ | 0.036 | 0.540 | 0.932 | 0.665 | 0.999 |
| TG_53:2_[M+NH4]1+ | 0.037 | 0.531 | 0.932 | 0.457 | 0.999 |
| PI-O_38:3_[M+NH4]1+ | 0.033 | 0.566 | 0.932 | 0.358 | 0.999 |
| TG_54:7_[M+NH4]1+ | 0.038 | 0.514 | 0.932 | 0.786 | 0.999 |
| TG_54:6_[M+NH4]1+ | 0.041 | 0.477 | 0.932 | 0.974 | 0.999 |
| TG_54:5_[M+NH4]1+ | 0.043 | 0.456 | 0.932 | 0.944 | 0.999 |
| TG_54:4_[M+NH4]1+ | 0.036 | 0.533 | 0.932 | 0.768 | 0.999 |
| TG_54:3_[M+NH4]1+ | 0.043 | 0.459 | 0.932 | 0.511 | 0.999 |
| PI_38:4_[M+NH4]1+ | -0.054 | 0.331 | 0.932 | 0.509 | 0.999 |
| TG_54:2_[M+NH4]1+ | 0.043 | 0.465 | 0.932 | 0.429 | 0.999 |
| TG_54:1_[M+NH4]1+ | 0.016 | 0.779 | 0.932 | 0.694 | 0.999 |
| TG_56:9_[M+NH4]1+ | 0.029 | 0.617 | 0.932 | 0.594 | 0.999 |
| TG_56:8_[M+NH4]1+ | 0.017 | 0.764 | 0.932 | 0.748 | 0.999 |
| TG_56:7_[M+NH4]1+ | 0.030 | 0.596 | 0.932 | 0.531 | 0.999 |
| TG_56:6_[M+NH4]1+ | 0.039 | 0.492 | 0.932 | 0.550 | 0.999 |
| TG_56:5_[M+NH4]1+ | 0.049 | 0.379 | 0.932 | 0.276 | 0.999 |
| TG_56:4_[M+NH4]1+ | 0.031 | 0.591 | 0.932 | 0.445 | 0.999 |
| PI_40:2_[M+NH4]1+ | 0.033 | 0.560 | 0.932 | 0.375 | 0.999 |
| TG_58:10_[M+NH4]1+ | 0.071 | 0.216 | 0.932 | 0.067 | 0.999 |
| TG_58:9_[M+NH4]1+ | 0.021 | 0.716 | 0.932 | 0.798 | 0.999 |

| | | | | | |
|---|---|---|---|---|---|
| TG_58:8_[M+NH4]1+ | 0.026 | 0.639 | 0.932 | 0.541 | 0.999 |
| TG_58:7_[M+NH4]1+ | 0.012 | 0.822 | 0.943 | 0.682 | 0.999 |

Supplementary Table 4. PVAR-systems GMM coefficients

| | WLZ | | LAZ | | WAZ | | ME1 | | ME2 | | ME3 | | ME4 | |
|---|---|---|---|---|---|---|---|---|---|---|---|---|---|---|
| | Coef | *p value* | Coef | *p value* | Coef | *p value* | Coef | *p value* | Coef | *p value* | Coef | *p value* | Coef | *p value* |
| constant | -0.342 | 0.000 | -0.308 | 0.000 | -0.410 | 0.000 | -0.002 | 0.603 | -0.003 | 0.245 | 0.002 | 0.581 | -0.002 | 0.503 |
| sex | -0.092 | 0.188 | 0.000 | 0.993 | -0.068 | 0.132 | 0.003 | 0.268 | -0.005 | 0.028 | -0.005 | 0.033 | -0.004 | 0.146 |
| WLZ t-1 | 0.149 | 0.151 | 0.304 | 0.008 | 0.072 | 0.402 | 0.008 | 0.139 | 0.002 | 0.696 | 0.006 | 0.235 | 0.002 | 0.728 |
| LAZ t-1 | -0.027 | 0.796 | 0.752 | 0.000 | 0.207 | 0.013 | 0.009 | 0.111 | 0.004 | 0.494 | 0.009 | 0.136 | -0.001 | 0.866 |
| WAZ t-1 | 0.399 | 0.017 | -0.112 | 0.529 | 0.506 | 0.000 | -0.014 | 0.096 | -0.006 | 0.495 | -0.010 | 0.223 | -0.001 | 0.920 |
| ME1 t-1 | 0.036 | 0.625 | 0.024 | 0.701 | 0.020 | 0.837 | 0.027 | 0.850 | 0.038 | 0.794 | 0.120 | 0.390 | 0.191 | 0.184 |
| ME2 t-1 | 0.190 | 0.000 | 0.188 | 0.000 | -0.236 | 0.000 | -0.047 | 0.537 | -0.029 | 0.674 | -0.095 | 0.240 | -0.161 | 0.047 |
| ME3 t-1 | 0.102 | 0.042 | 0.125 | 0.005 | -0.115 | 0.053 | 0.019 | 0.868 | 0.065 | 0.627 | 0.119 | 0.351 | 0.141 | 0.121 |
| ME4 t-1 | 0.051 | 0.215 | 0.151 | 0.000 | -0.092 | 0.121 | 0.010 | 0.916 | -0.036 | 0.684 | 0.074 | 0.485 | -0.079 | 0.455 |
| ME5 t-1 | 0.019 | 0.752 | -0.032 | 0.489 | -0.014 | 0.862 | 0.040 | 0.751 | -0.051 | 0.653 | 0.216 | 0.069 | -0.160 | 0.296 |
| ME6 t-1 | 0.161 | 0.000 | 0.162 | 0.000 | -0.211 | 0.001 | 0.087 | 0.128 | -0.054 | 0.328 | -0.088 | 0.153 | -0.020 | 0.765 |
| ME7 t-1 | -0.118 | 0.132 | 0.020 | 0.776 | 0.133 | 0.232 | -0.115 | 0.380 | 0.150 | 0.262 | -0.064 | 0.597 | -0.333 | 0.020 |
| ME8 t-1 | 0.034 | 0.562 | 0.143 | 0.001 | -0.055 | 0.414 | 0.125 | 0.285 | 0.091 | 0.390 | -0.001 | 0.992 | 0.173 | 0.131 |
| ME9 t-1 | 0.216 | 0.004 | 0.225 | 0.000 | -0.236 | 0.017 | 0.148 | 0.308 | 0.053 | 0.640 | -0.097 | 0.467 | -0.014 | 0.912 |
| ME10 t-1 | 0.139 | 0.001 | 0.142 | 0.000 | -0.171 | 0.003 | -0.078 | 0.364 | -0.055 | 0.534 | 0.082 | 0.354 | -0.018 | 0.813 |
| ME11 t-1 | 0.107 | 0.008 | 0.145 | 0.000 | -0.146 | 0.006 | 0.118 | 0.339 | -0.056 | 0.430 | 0.021 | 0.847 | 0.216 | 0.064 |

red text denotes significant at 5% level

Supplementary Table 4. PVAR-systems GMM coefficients

| | ME5 | | ME6 | | ME7 | | ME8 | | ME9 | | ME10 | | ME11 | |
|---|---|---|---|---|---|---|---|---|---|---|---|---|---|---|
| | Coef | *p value* | Coef | *p value* | Coef | *p value* | Coef | *p value* | Coef | *p value* | Coef | *p value* | Coef | *p value* |
| constant | -0.004 | 0.211 | -0.004 | 0.117 | -0.007 | 0.048 | -0.009 | 0.005 | -0.002 | 0.561 | -0.009 | 0.005 | -0.010 | 0.001 |
| sex | -0.004 | 0.100 | -0.007 | 0.003 | 0.002 | 0.423 | 0.000 | 0.824 | -0.004 | 0.120 | -0.003 | 0.159 | -0.004 | 0.067 |
| WLZ t-1 | -0.007 | 0.268 | -0.016 | 0.006 | 0.027 | 0.000 | 0.014 | 0.017 | -0.007 | 0.250 | 0.005 | 0.378 | 0.005 | 0.396 |
| LAZ t-1 | -0.006 | 0.355 | -0.015 | 0.014 | 0.031 | 0.000 | 0.018 | 0.004 | -0.005 | 0.439 | 0.003 | 0.600 | 0.005 | 0.379 |
| WAZ t-1 | 0.003 | 0.777 | 0.022 | 0.013 | -0.047 | 0.000 | -0.028 | 0.001 | 0.006 | 0.475 | -0.008 | 0.357 | -0.012 | 0.164 |
| ME1 t-1 | -0.096 | 0.547 | -0.004 | 0.974 | -0.042 | 0.749 | 0.026 | 0.880 | 0.379 | 0.002 | 0.016 | 0.918 | -0.452 | 0.002 |
| ME2 t-1 | 0.000 | 0.998 | -0.136 | 0.078 | 0.131 | 0.073 | 0.020 | 0.809 | 0.171 | 0.056 | 0.124 | 0.178 | -0.025 | 0.714 |
| ME3 t-1 | -0.380 | 0.002 | -0.292 | 0.013 | 0.102 | 0.397 | 0.040 | 0.681 | 0.068 | 0.533 | 0.344 | 0.001 | 0.123 | 0.254 |
| ME4 t-1 | 0.139 | 0.173 | 0.299 | 0.001 | 0.136 | 0.115 | 0.037 | 0.724 | -0.126 | 0.274 | -0.171 | 0.168 | -0.028 | 0.767 |
| ME5 t-1 | -0.024 | 0.841 | 0.004 | 0.978 | -0.090 | 0.329 | -0.082 | 0.423 | 0.230 | 0.043 | -0.053 | 0.590 | -0.129 | 0.293 |
| ME6 t-1 | 0.043 | 0.464 | -0.021 | 0.723 | 0.109 | 0.098 | 0.001 | 0.988 | -0.057 | 0.285 | 0.053 | 0.420 | 0.024 | 0.674 |
| ME7 t-1 | -0.082 | 0.537 | 0.389 | 0.001 | 0.051 | 0.702 | 0.275 | 0.054 | 0.050 | 0.708 | 0.063 | 0.622 | 0.486 | 0.000 |
| ME8 t-1 | 0.062 | 0.627 | 0.282 | 0.008 | 0.135 | 0.167 | 0.201 | 0.076 | 0.067 | 0.553 | -0.048 | 0.658 | -0.007 | 0.946 |
| ME9 t-1 | 0.128 | 0.332 | -0.033 | 0.773 | -0.067 | 0.553 | 0.061 | 0.602 | 0.403 | 0.000 | -0.200 | 0.064 | -0.391 | 0.000 |
| ME10 t-1 | -0.121 | 0.170 | -0.077 | 0.353 | 0.247 | 0.006 | -0.035 | 0.699 | 0.086 | 0.324 | 0.225 | 0.041 | -0.197 | 0.005 |
| ME11 t-1 | 0.096 | 0.397 | 0.084 | 0.408 | 0.116 | 0.136 | 0.001 | 0.986 | -0.228 | 0.005 | 0.103 | 0.254 | 0.078 | 0.487 |

red text denotes significant at 5% level

Supplementary Table 5. Longitudinal metabolomics or lipidomics studies among children in early years of life

| Study | Population | Analysis | Platform | Data analysis (main) | Findings |
|---|---|---|---|---|---|
| Chiu et al [1] | Taiwan<br>Age: 1-4 years old<br>Sampled at 6 months, 1, 2, 3 and 4 years | Urine metabolomics | $^{1}H$ NMR | Partial least squares-based model | Higher urinary trimethylamine N-oxide (TMAO) and betaine level in children aged 6 months. Glycine and glutamine levels declined after 6 months with an increase in creatine and creatinine. Pathways associated with amino acid metabolism were different between infants aged 6 months and 1 year, whereas pathways associated with carbohydrate metabolism were different between children at ages 2 and 3 years. |
| Chiu et al [2] | Taiwan<br>Age: 1-4 years old<br>Sampled at 6 months, 1, 2, 3 and 4 years | Urine metabolomics | $^{1}H$ NMR | Partial least squares-based model | Urinary metabolites are associated to the development of asthma in children |
| Neyraud et al[3] | France<br>Age: 3 – 11 months<br>Sampled at 3, ~4.5, ~5.5 and 11th month | Saliva metabolomics | $^{1}H$ NMR | Partial least squares-based model<br>ANOVA – simultaneous component analysis | Significant changes in saliva metabolome over time but was independent of milk feeding history |
| Giallourou et al[4] | Peru, Bangladesh, Tanzania<br>Age: 3 – 24 months<br>Sampled at 3, 6, 9, 15 and 24 months for urine; 7 and 15 months for plasma | Urine and plasma metabolomics | $^{1}H$ NMR and LC-MS | Partial least squares-based model | Developed a phenome-for-age Z score (PAZ) as a measure of metabolic maturity. Growth restricted children lagged in terms of PAZ and that PAZ is predictive of length-for-age z score (a measure of linear growth). |
| Nikkilä et al[6] | Finland<br>Age: birth to 2 years | Serum lipidomics | LC-MS | Hidden Markov chains | Levels of sphingomyelins were higher in girls compared to boys in all 5 hidden Markov metabolic states, |

| | | | | | |
|---|---|---|---|---|---|
| | Sampled at 3 month intervals | | | | indicated marked sex-based differences in lipidome progression in infants |
